# Practical barriers and facilitators experienced by patients, pharmacists and physicians to the implementation of pharmacogenomic screening in Dutch outpatient hospital care – an explorative pilot study

**DOI:** 10.1101/2020.11.11.20229211

**Authors:** P. Lanting, W.H. Drenth, L.G. Boven, A. van Hoek, A.M.A. Hijlkema, A.E. Poot, G. van der Vries, R.A. Schoevers, R.O.B. Gans, J.G.W. Kosterink, M. Plantinga, I.M. van Langen, A.V. Ranchor, C. Wijmenga, L.H. Franke, B. Wilffert, R.H. Sijmons

**Affiliations:** University of Groningen, University Medical Center Groningen, Department of Genetics, Groningen, The Netherlands; University of Groningen, University Medical Center Groningen, Department of Psychiatry, Groningen, The Netherlands; University of Groningen, University Medical Center Groningen, Department of Internal Medicine, Groningen, The Netherlands; University of Groningen, University Medical Center Groningen, Department of Clinical Pharmacy and Pharmacology, Groningen, The Netherlands; University of Groningen, Groningen Research Institute of Pharmacy, Unit of PharmacoTherapy, -Epidemiology & -Economics, Groningen, The Netherlands; University of Groningen, University Medical Center Groningen, Department of Health Psychology, Groningen, The Netherlands

**Keywords:** Pharmacogenetics, pharmacogenetic, pharmacogenomics, pharmacogenomic, implementation, screening, pre-emptive, personalized medicine, precision medicine

## Abstract

Pharmacogenomics (PGx) can provide optimized treatment to individual patients while potentially reducing healthcare costs. However, widespread implementation remains absent. We performed a pilot study of PGx screening in Dutch outpatient hospital care to identify the barriers and facilitators to implementation experienced by patients (n=165), pharmacists (n=58) and physicians (n=21). Our results indeed suggest that the current practical experience of healthcare practitioners (HCPs) with PGx is limited, that proper education is necessary, that patients want to know the exact implications of the results, and that there is an unclear allocation of responsibilities between HCPs about who should discuss PGx with patients and apply PGx results in healthcare. We observed a positive attitude toward PGx among all the stakeholders in our study, and among patients this was independent of the occurrence of drug-gene interactions during their treatment. Facilitators included the availability of and adherence to Dutch Pharmacogenetic Working Group guidelines. While Clinical Decision Support (CDS) is available and valued in our medical center, the lack of availability of CDS might be an important barrier within Dutch healthcare in general.

## INTRODUCTION

Pharmacogenomics (PGx) can help predict which medication will be most effective and safe in individual patients while potentially reducing healthcare costs.^1,2^ Ideally, an individual’s PGx profile would be known before drug prescription ‒ an approach known as pre-emptive PGx testing or PGx screening ‒ rather than being determined after observing low therapeutic response or adverse drug reactions (ADRs). Potential benefits of introducing PGx screening into a routine healthcare setting include reduced hospitalizations and cost, and improved safety, adherence and efficacy.^3^ Dutch national guidelines on practical application of PGx for drug prescription developed by the Dutch Pharmacogenetics Working Group (DPWG) are available through the Dutch drug database, referred to as the G-standard.^4,5^ Based on these DPWG guidelines, it is estimated that an alternative dosage or drug would be recommended for 1 in 20 drug prescriptions in primary care if PGx screening became the standard-of-care in the Netherlands.^6^ Nevertheless, PGx is rarely applied in current clinical practice.^2,7^

A number of barriers to PGx implementation have been identified so far. These include unclear procedures, insufficient evidence, inefficient infrastructure, lack of a standardized format for reporting results, lack of ICT support tools, and lack of knowledge, training and experience among healthcare practitioners (HCPs). Reported facilitators include recognition of clinical utility, pharmacist’s feelings of responsibility for delivering PGx to patients, and the availability of professional guidelines for interpreting test results.^1,8,9,10,11,12,13^ To the best of our knowledge, no study has identified barriers and facilitators from the perspective of all the relevant stakeholders in an actual implementation setting. Therefore, we carried out an explorative pilot study to identify such barriers and facilitators while offering PGx screening in two outpatient clinics of the University Medical Center of Groningen (UMCG) in the Netherlands.

## METHODS

This study was designed as an explorative pilot study with mixed methods. The study timeline is shown in **Figure 1A** and an overview of the study design in **Figure 1B**. Additional background information is provided in **Supplementary Methods** section 1.

**Figure 1:**
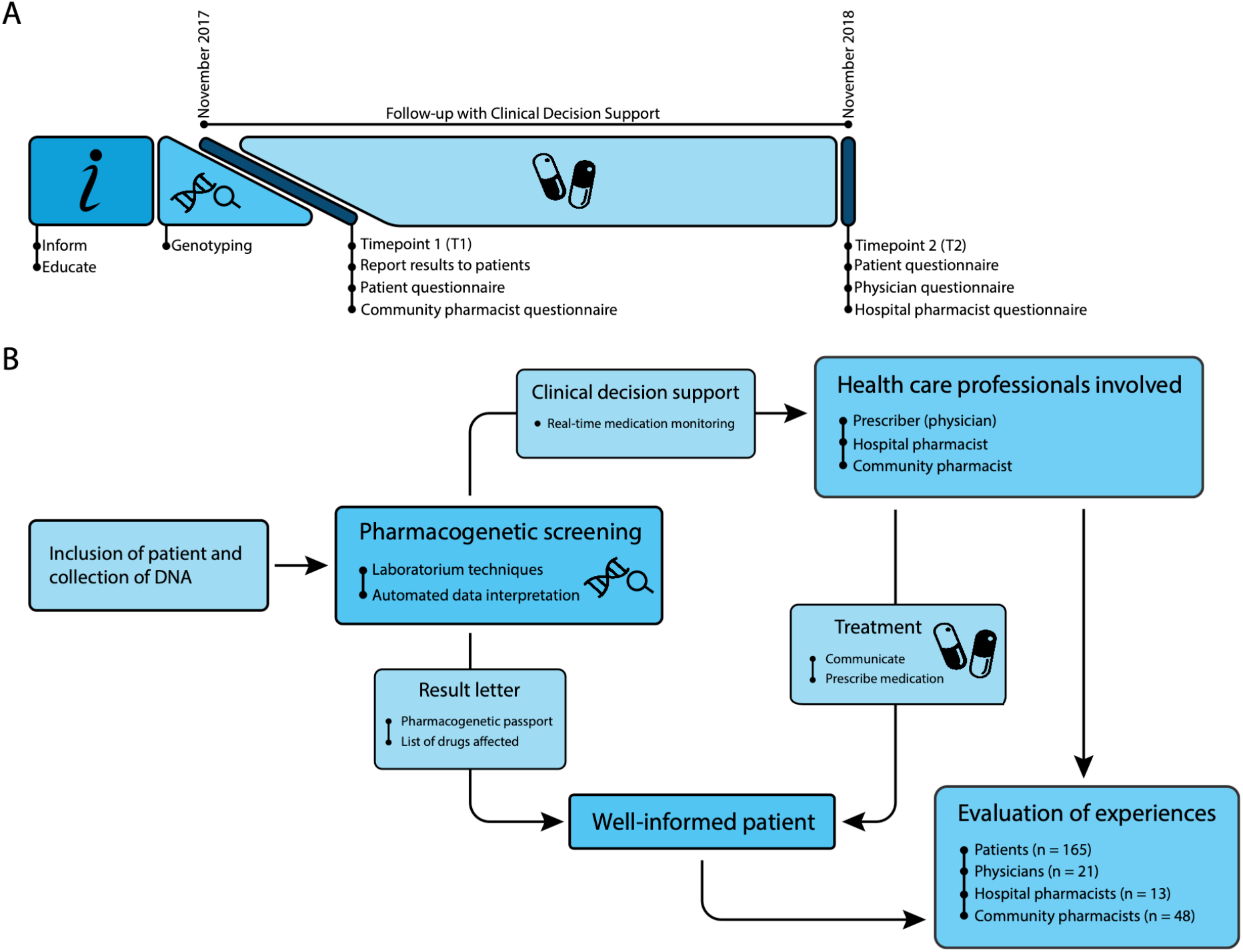
Study timeline and design. [**A**. Study timeline **B**. Study design]

### Recruitment of participants

The outpatient clinics of Internal Medicine and Psychiatry, and the hospital pharmacy of the UMCG were approached to participate in this study. Information about the study’s aim was provided during an introductory meeting with each department. Physicians who took part recruited participants from their own patients on a first-come-first-served basis until the study test capacity of 165 PGx individuals was reached. Inclusion criteria were: 18 years or older, cognitively competent, able to read and speak Dutch, and able to provide a blood sample. Eligible patients received printed information about the project goal, procedures for testing, reporting of results and links to resources with additional information (project website and animated video). See **Supplementary Materials 1-4**.

Community pharmacists listed in the patient’s Electronic Health Record (EHR) were invited to fill out questionnaires by mail simultaneously with the reporting of PGx screening results (T1). UMCG physicians at the two clinics and hospital pharmacists involved in patient care were invited to fill out study questionnaires by email at the end of follow-up (T2, **Figure 1A**). See **Supplementary Methods** section 3 for additional details.

### Genotyping and reporting of PGx screening results

After providing written informed consent, patients underwent genotyping with a custom panel of 14 genes (**Table S1**). Next, patients received a letter with their PGx screening results and an explanation in layman’s terms (see **Supplementary Materials 5**). Copies were also stored in their hospital EHR and sent to their community pharmacist and general practitioner (GP) (**Figure 1B**). See **Supplementary Methods** section 4 for additional details.

Custom Clinical Decision Support (CDS) software developed prior to the study was used to provide hospital prescribers with relevant DPWG recommendations in real-time during drug prescription (**Figure 1**). See **Supplementary Methods** section 5 for additional details.

### Data collection

PGx screening results, predicted drug-gene interactions (DGIs), and CDS use including user comments and actions taken based on recommendations were stored in the study database. Relevant medical information, including patient drug use during the follow-up period November 2017–November 2018, was manually extracted from EHRs (see **Supplementary Methods** section 6 for additional details). Follow-up started from the time the results were reported and therefore varied between patients, up to a maximum of a year (**Figure 1A**). We conducted five questionnaires to evaluate the experiences of patients, physicians, and pharmacists via open- and closed-ended questions at time of result reporting (T1) and after follow-up (T2, **Figure 1A**). The survey study was designed by a multidisciplinary team using input from an explorative qualitative interview and focus group study with 13 prescribers from the participating outpatient clinics, 13 patients and 7 pharmacists (see **Supplementary Methods** section 2). The questionnaires included items on various themes: socio-demographics, knowledge and education about PGx, attitude towards PGx screening, application of PGx, provision of information about PGx, and result reporting (**Table S2**). The attitude questions originate from the Theory of Planned Behavior Framework.^14^ All other questions were self-constructed. The two patient questionnaires were sent out on paper, with the option to respond digitally, at the time of results reporting (T1) and after follow-up (T2). If necessary, patients were reminded by mail and again by telephone to respond. Community pharmacists were invited to respond to the survey on paper, with the option to respond digitally, at the time results were reported (T1). The outpatient clinic physicians and hospital pharmacists received an invitation for a digital survey by email after follow-up (T2, **Figure 1A**). Digital survey responses were collected using the routine outcome monitoring application RoQua.^15^ Responses on paper were registered in RoQua by the researchers.

### Data analysis

CDS searches and survey responses to open-ended questions were independently categorized by two researchers (AvH, AMAH), and discrepancies were resolved by an independent third researcher (PL). All data collected was pseudonymized and analyzed per theme using R.^16^ For survey responses, the Shapiro-Wilk test was used to assess normality. Subsequent subgroup comparisons were performed using a *t*-test or Wilcoxon test. Cronbach’s alfa was used to assess internal consistency of survey questions.

### Ethical approval

This study was approved by the Medical Ethics Review Board of the UMCG (reference: 2017.266).

## RESULTS

### Participants

This study included 165 patients, 21 physicians, 13 hospital pharmacists, and 48 community pharmacists (**Figure 1B**) and explored various themes around practical barriers and facilitators. Response rates to the patient questionnaires were 84% (n=138, T1) and 74% (n=122, T2). Response rates to the HCP questionnaires were 19% (physicians, T2), 28% (hospital pharmacists, T2), and 77% (community pharmacists, T1). Response rates per survey item varied since not all respondents have answered all items. Median patient follow-up was 244 days (range: 117-365). See **Table S3** for full demographics of study participants.

### Screening results, drug use and DGIs

Out of the study population, 158 patients (96%) carried at least one actionable PGx haplotype or predicted PGx phenotype (**Table S4** lists frequencies of PGx haplotypes and predicted PGx phenotypes). During follow-up, 60 patients received drug treatment (36%). Following DPWG guidelines, DGIs were observed in 21 patients (13%): 18 with one DGI, one with two DGIs and two with three DGIs. Actionable DGIs were observed in 20 patients (12%): 18 with one actionable DGI and two with two actionable DGIs. In total, 120 unique drugs were used during follow-up, including 18 with a known DGI (15%), of which 15 were actionable in the study population. During follow-up, patients used two drugs (range: 0–13 drugs) on average, and 27 patients (23% of T2 respondents) reported being prescribed at least one new drug. Patients reported that prescriptions originated from their GP’s office (83% of T2 respondents) or hospital physician (17% of T2 respondents). See **Supplementary Results** section 1 for survey results on the review of patient drug use in response to PGx screening results.

### CDS searches and output during follow-up

During follow-up, CDS was used to consult the DPWG guidelines 59 times for 20 patients. A CDS search was performed for eight patients who received drug treatment, and four had DGIs. CDS searches were performed by prescribers from participating outpatient clinics, who were explicitly instructed to use CDS to consult DPWG guidelines, and other prescribers in the hospital, who were either informed about the PGx screening results by the patient or encountered the results in the EHR. CDS searches were categorized into six subgroups using treatment information from the EHR: prescribing situation (5%), cascade (search in response to previous search) (2%), potential future treatment (29%), current treatment (47%), past treatment (5%), and other (12%) (**Figure 2A**).

**Figure 2:**
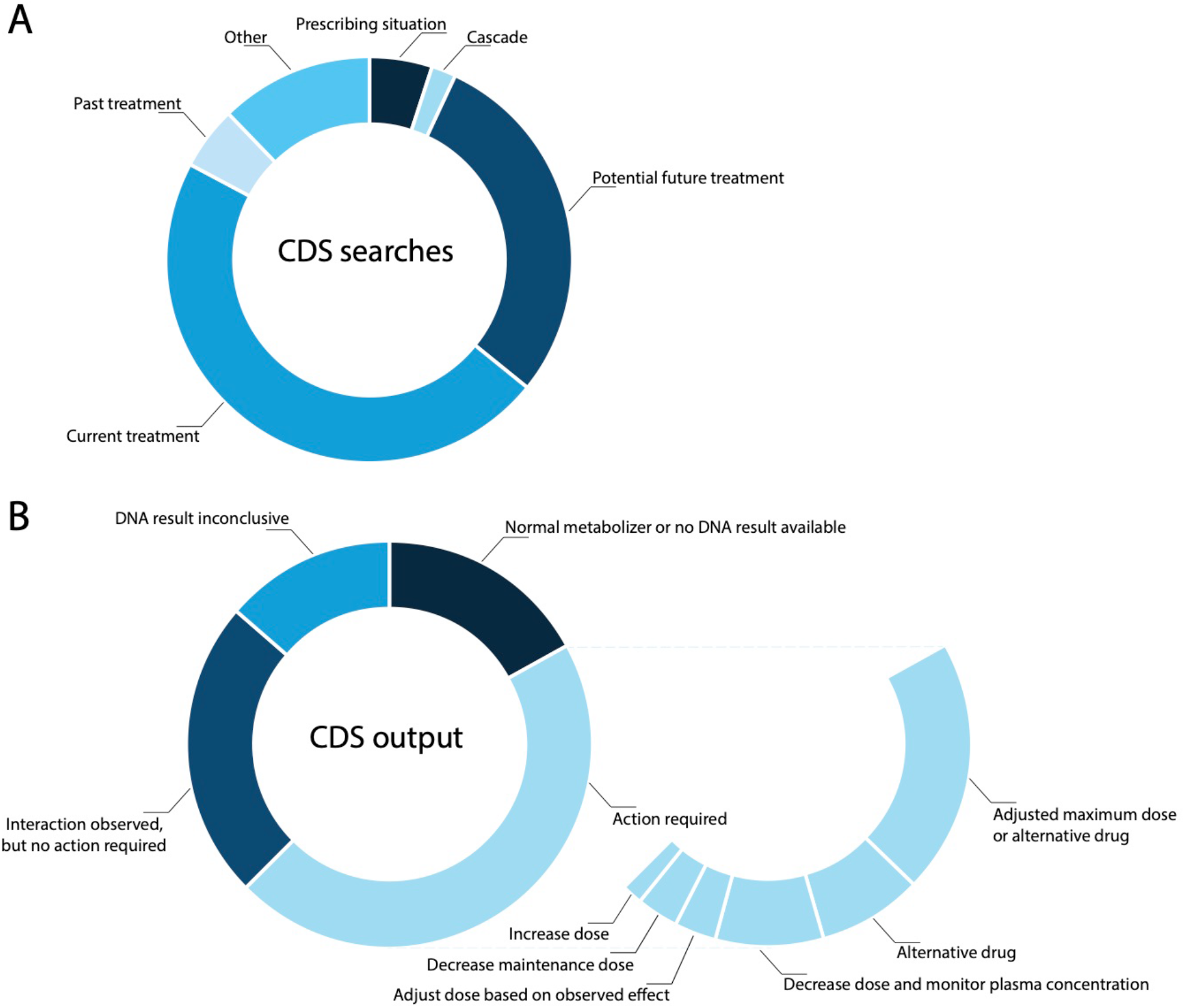
CDS searches and output. [**A**. Searches performed by physicians in the Clinical Decision Support software **B**. Output provided to the physician by the Clinical Decision Support software]

Of the CDS searches, 27 (45.8%) yielded recommendations requiring an action by the prescriber, 14 (23.7%) did not require an action, 10 (16.9%) found no available recommendations (e.g. in case of normal metabolizers), and 8 (13.6%) had inconclusive DNA test results. Of the actionable recommendations, 12 (44%) advised adhering to an adjusted maximum (daily) dose or prescribing an alternative, 5 (19%) advised prescribing an alternative, 5 (19%) advised lowering the dose and monitoring plasma concentrations, 2 (7%) advised adjusting the dose based on the effect observed, 2 (7%) advised lowering the maintenance dose and 1 (4%) advised increasing the dose (**Figure 2B**). Details of the DGIs involved and an evaluation of DPWG guidelines are presented in **Supplementary Results** sections 2 and 3.

### Prior experience of HCPs with PGx

Twenty-one community pharmacists (44%), one hospital pharmacist (8%), and five physicians (24%) reported that this study was their first experience with PGx test results. One in eight community pharmacists, six hospital pharmacists (46%) and half of physicians (52%) reported having taken the initiative to conduct PGx testing at least once in the past. These results highlight that current practical experience is limited.

### Knowledge and education of HCPs

In all professions, half the HCPs reported having received postgraduate education about PGx. The self-graded knowledge level was significantly higher in these subgroups (**Table 1**). Pharmacists reported a need for further education, both for themselves (n=47, 77%) and for pharmacy staff (n=52, 87%), whereas physicians did not report this need.

**Table 1.** Self-graded knowledge and application level of HCPs.

| Healthcare practitioner |  | Self-graded knowledge level |  | Self-graded application level |  |
| --- | --- | --- | --- | --- | --- |
| Community pharmacists | with postgraduate education | 6.5 (4–8) | p = 0.011 | 7 (5–10) | p = 0.005 |
|  | without postgraduate education | 6 (2–7) |  | 5 (2–8) |  |
| Hospital pharmacists | with postgraduate education | 7.7 (7–9) | p = 0.01 | 7.5 (7–9) | p = 0.016 |
|  | without postgraduate education | 6.3 (5–7) |  | 6 (2–7) |  |
| Physicians | with postgraduate education | 7 (1–9) | p = 0.002 | 7 (6–8) | p = 0.203 |
|  | without postgraduate education | 4 (6–8) |  | 6 (3–9) |  |

### Patient attitudes towards PGx screening after follow-up (T2)

Most patients reported that genetic testing in general (n=89, 77%) or PGx testing (n=102, 88%) did not frighten them. Knowing their PGx profile was considered comforting (n=106, 89%) and useful (n=111, 92%), and patients thought that it has added value when their pharmacotherapy is adjusted using PGx (n=107, 91%). No significant difference was found in the attitude of patients with or without observed DGIs.

### HCP attitudes towards PGx screening

Nearly all HCPs were positive about the usefulness of PGx information for their patients (useful to have: n=69, 84%; would like to use more in daily practice: n=72, 88%; added value: n=71, 87%). However, nine community pharmacists (19%), two hospital pharmacists (15%) and four physicians (20%) did not feel ready to apply PGx information in daily practice.

### Practical application of PGx

Community pharmacists graded their expected application level (T1), whereas hospital pharmacists and physicians graded their perceived application level (T2). The self-graded application level is significantly higher in the education subgroups for both community and hospital pharmacists, but not for physicians (**Table 1**). Prominent arguments provided to explain higher self-graded application levels were that application of PGx was possible with use of the pharmacy or hospital computer system (n=12) and that HCPs had come across PGx more often (during education or in practice) (n=8). Notable arguments to explain lower self-graded application levels were that HCPs perceived insufficient knowledge themselves (n=8) and reported practical barriers present within computer systems, for example that not all PGx results could be registered (n=5). In summary, HCPs relied heavily on their computer system for the application of PGx, perceived a need for education on PGx application, and experienced practical barriers within computer systems that hindered PGx application. **Supplementary Results** section 4 describes an event that occurred during follow-up that illustrates the importance of educating and informing all HCPs involved in practical application of PGx.

### Patients’ needs for information about their PGx screening results

After receiving the PGx screening results (T1), 15 patients (11%) reported still having questions with respect to these results, most often wanting to know the exact implications, e.g. the level of dose adjustment or suitable alternative drugs (n=6). Patients generally consulted their treating physician in the hospital during follow-up to gain additional information. After follow-up (T2), the number of patients having questions about their PGx screening results has increased to 23 (19%). They still primarily wanted to know the implications of the results for them (n=7). Thirty-six patients (30%) reported that improvements could be made in the information provided, most importantly in explaining the exact implications of the results for them (n=9), providing better explanation in general (n=7), and better educating HCPs (n=4). Detailed evaluation of the PGx result letter is presented in **Supplementary Results** section 5. In summary, some patients wished to receive more and different information than provided in this study.

### Discussing PGx screening results with patients: patient surveys

After receiving the PGx screening results (T1), 47 patients (35%) believed an HCP should always discuss these results with them, 29 (21%) only if patients express the need, and 33 (24%) only if the results have consequences. Twenty-six (19%) thought the results should not be discussed with them at all. According to patients, the preferred HCPs to discuss PGx screening results are the treating physician in the hospital (n=80, 44%), GP (n=47, 26%), clinical geneticist (n=30, 16%), or pharmacist (n=22, 12%).

After receiving the PGx screening results (T1), 101 patients (74%) planned to discuss them with their treating physician, with 44 patients (37%) reporting having done so after follow-up (T2) in a regular appointment and 6 (5%) reporting having done so in a separate appointment. In total, 101 conversations about PGx screening results between patients and HCPs were scored by patients (46% physician, 21% community pharmacist, 21% GP, 8% physician from other hospital, 2% home nurse, 2% thrombosis care, and 1% nursing home). Seventy-one percent of these conversations were scored as ‘(very) good’. In one case, the conversation was scored as ‘good’, but the patient reported that the HCP did not (fully) understand the results. Thirteen percent of conversations were scored as ‘(very) bad’. In two cases, the conversation as such was scored as ‘(very) bad’ even though, on a positive note, the HCP had started using the PGx results **(Figure 3)**.

**Figure 3:**
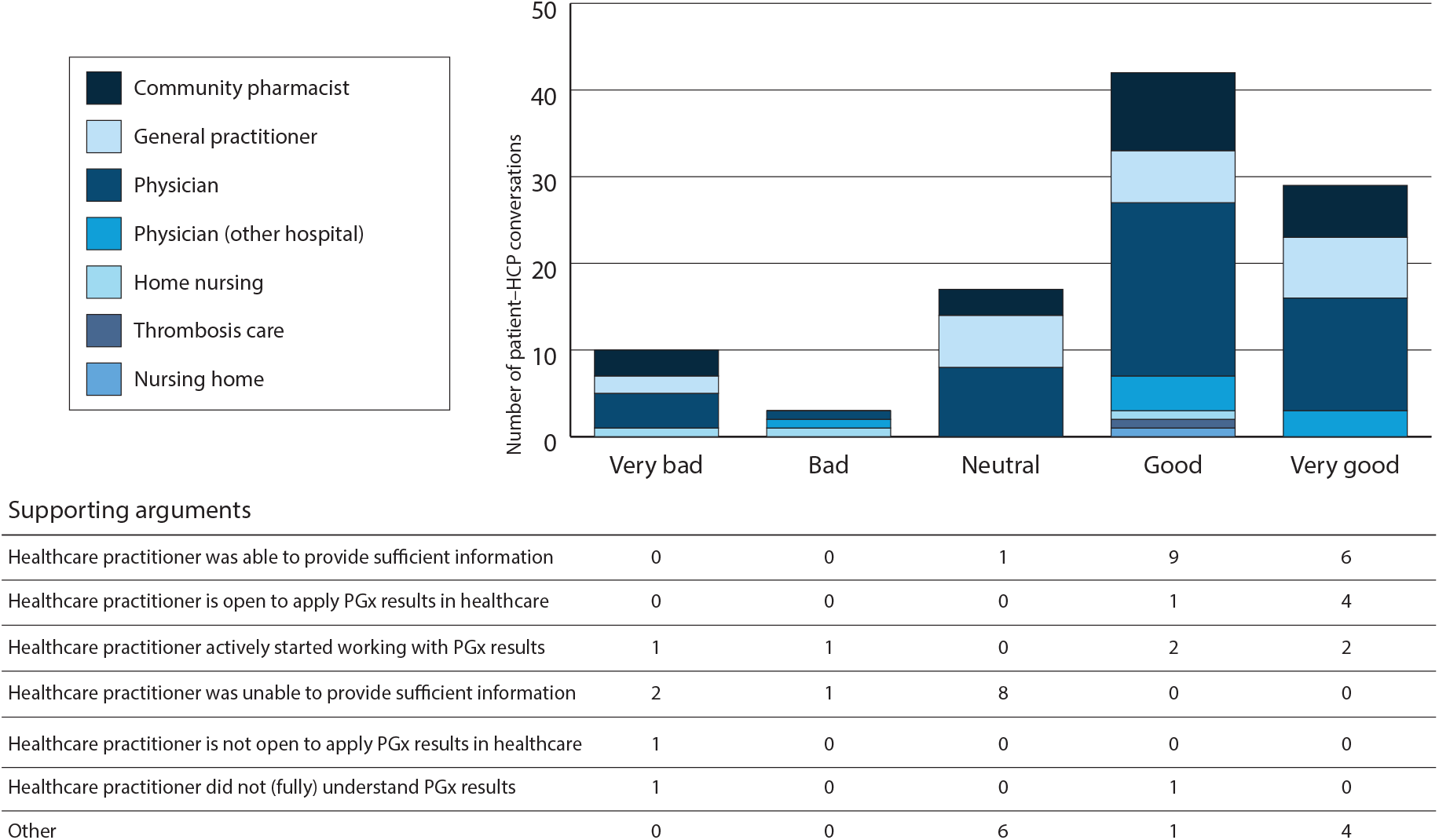
Conversation scores for the discussion of PGx test results with HCPs. [The number of conversations between patients and different HCPs, the score patients gave to those conversations, and the supporting arguments for the score given.]

### Discussing PGx screening results with patients: HCP surveys

Sixteen community pharmacists (36%), eight hospital pharmacists (62%) and 13 physicians (62%) believed that PGx screening results should always be discussed with patients by an HCP, with eight (18%), two (15%) and five (24%), respectively, believing it should only be done if a patient expresses the need and 19 (42%), three (23%) and three (14%), respectively, only if the results have consequences. Two community pharmacists (4%) did not believe results should be discussed with patients at all. Community pharmacists primarily placed the responsibility for discussing PGx screening results with patients in the hands of the treating physician in the hospital (n=26, 38%) or pharmacist (n=21, 31%), and to a lesser extent with the clinical geneticist (n=13, 19%). Hospital pharmacists also primarily placed this responsibility in the hands of the treating physician in the hospital (n=11, 39%) or pharmacist (n=8, 29%), and to a lesser extent with the GP (n=4, 14%) or clinical geneticist (n=4, 14%). Physicians primarily indicated that they, as treating physicians in the hospital, should discuss PGx screening results with patients (n=19, 59%), followed by the pharmacist (n=7, 22%) and the GP (n=3, 9%).

Community pharmacists were asked what they planned to do with the PGx screening results they had received (T1). All plans reported for PGx screening results are shown in **Figure 4**. Although four community pharmacists reported that PGx screening results should always be discussed with the patient by an HCP, preferably the pharmacist, none of these four pharmacists reported that they themselves intended to discuss the results with their patients.

**Figure 4:**
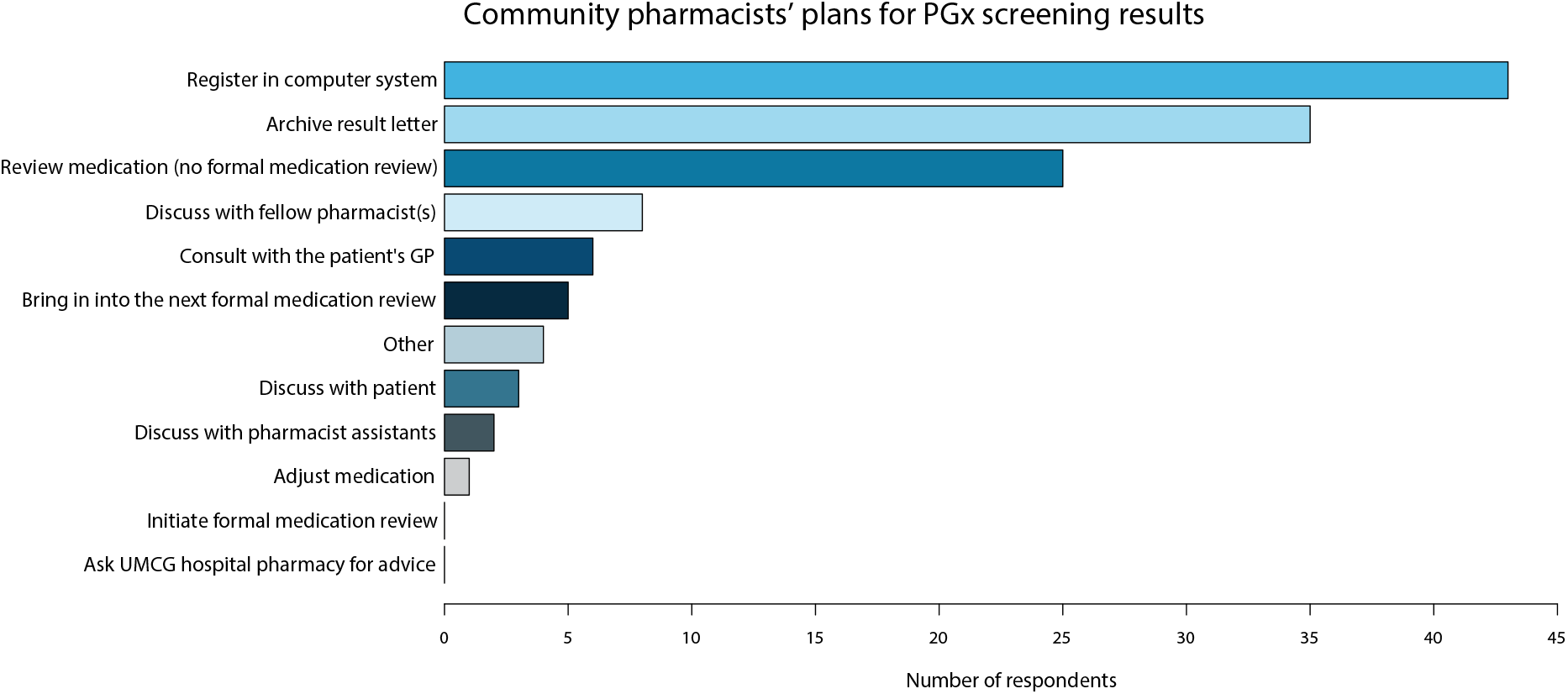
Community pharmacists’ plans for PGx screening results. [The steps which community pharmacists reported they would take after having received PGx screening results]

Five out of six hospital pharmacists and all eight physicians who discussed PGx screening results with patients and/or other HCPs felt they had sufficient knowledge to do so. None of them reported questions about PGx that they were unable to answer.

### Responsibility for application of PGx screening results in patient care

HCPs were also asked about who they regarded as having the final responsibility for the application of PGx screening results in patient care. The results are presented in **Table 2** and show that the majority of physicians reported that this responsibility lies with the prescriber. Hospital pharmacists largely agreed with this, although a notable group also reported the pharmacist as responsible. Community pharmacists were more divided and specifically indicated that there is a shared responsibility. In summary, the allocation of responsibility for the application of PGx screening results in patient care is currently unclear.

**Table 2:** Final responsibility for the application of PGx screening results in patient care.

| Responsible person | Community pharmacists | Hospital pharmacists | Physicians |
| --- | --- | --- | --- |
| Pharmacist | 18 (39%) | 5 (38%) | 2 (9.5%) |
| Prescriber | 10 (22%) | 8 (62%) | 16 (76%) |
| Clinical geneticist | 7 (15%) | - | 2 (9.5%) |
| General practitioner | - | - | - |
| Other |  | - |  |
| Shared responsibility in general | 5 (11%) |  |  |
| Pharmacist and prescriber are jointly responsible | 4 (9%) |  |  |
| Pharmacist, providing sufficient information transfer | 1 (2%) |  |  |
| Depending on drug prescribed | - |  | 1 (5%) |
| Other | 1 (2%) |  |  |

### Identified practical barriers and facilitators

An overview of the identified practical barriers and facilitators within the various themes discussed above, as perceived by HCPs and patients, is presented in **Table 3**.

**Table 3:** Barriers and facilitators to PGx implementation.

|  | Perceived by stakeholder |  |  |  |
| --- | --- | --- | --- | --- |
|  | Patient | Community pharmacist | Hospital pharmacist | Physician |
| **Barriers** |  |  |  |  |
| Practical experience is limited | No | Yes | Yes | Yes |
| Need for further postgraduate education | No | Yes | Yes | No |
| Rely on computer system for application | No | Yes | Yes | Yes |
| Need for education about PGx application | No | Yes | Yes | Yes |
| Practical barriers within computer systems | No | Yes | Yes | Yes |
| Lack of information, specifically about exact implications of PGx screening results | Yes | No | No | No |
| Unclear allocation of responsibilities among HCPs | Yes | Yes | Yes | Yes |
| **Facilitators** |  |  |  |  |
| Positive attitude towards PGx | Yes | Yes | Yes | Yes |

## DISCUSSION

This study identified practical barriers and facilitators within various themes, as perceived by HCPs and patients, to the use of PGx screening results and associated DPWG recommendations in a Dutch outpatient hospital care setting (**Table 3**). As some of the survey questions dealt with the actual outcome of PGx testing, we discuss these first.

### Frequencies of PGx variants and DGIs

We confirmed that actionable PGx variants are present in the majority of the patient population of outpatient clinics in frequencies comparable to those reported in literature (**Table S4**). To the best of our knowledge, this is the first study to report the number of DGIs after PGx screening where the screening was not initiated by the prescription of a drug with a known DGI. The median number of drugs used by participating patients during follow-up was two, whereas the median number of drugs dispensed to a patient in the same Northern Netherlands regions, as registered in the IADB.nl database, was three.^17^ We also only analyzed drugs recorded in the EHR. Since the majority of new prescriptions during follow-up originated from the GP, and drugs prescribed by GPs were not considered in our study, the number of DGIs we report is likely an underestimation. It is important that the number of DGIs is determined in more detail for a variety of patient populations in order to assess the value of PGx for individual patients.

CDS searches were performed in only four patients with a DGI, but recommendations were shown for more patients. This is explained by the fact that an alternative drug without a DGI was prescribed following the recommendation shown or because drugs were not prescribed directly following the search. The latter is illustrated by the search types we could distinguish. Some searches concerned past or future treatment, and prescribers also checked drugs they did not want to prescribe at that moment, for example commonly used treatment alternatives or drugs that were suggested in a recommendation. Furthermore, it is likely that prescribers started to remember the recommendations for DGIs they had encountered previously and did not perform a CDS search every time. The number of CDS searches reported is therefore likely to be an underestimation of the actual number of times prescribers dealt with PGx results. Although a comparison to standard-of-care is not possible based on available data, and we cannot discriminate between the different reasons for the changes made after drug review during this study, it is likely that DPWG recommendations altered prescription choices. From the actionable recommendations evaluated, we conclude that DPWG guidelines are generally well adhered to, although practical application can transcend guideline recommendations and application is thus not always straightforward.

### Practical barriers and facilitators

In agreement with literature, our results show that current practical experience with PGx is limited, even though DPWG guidelines have been available nationwide since 2006.^2,4,7^ A lack of knowledge and training amongst HCPs has previously been reported as a barrier to PGx implementation.^1,8,9,10,12,13^ The community and hospital pharmacists in our study reported wanting more education about PGx for themselves and pharmacy staff. Physicians in our study did not report this, which does not directly imply that they have enough knowledge or skills given that some also reported not feeling ready to apply PGx in daily practice. While physicians themselves perceived the general introduction and presentation of DPWG guideline recommendations provided in this study as sufficient, some patients wanted physicians to be better informed. According to these patients, some physicians were unable to provide sufficient explanation or did not fully understand the results. Our findings suggest that postgraduate education could increase the ability of HCPs to apply PGx in practice. Due to the explorative nature of our study, we can only speculate that the currently available training may not correspond well with practical needs (specifically on the topic of communication with patients), that training may not be optimized for physicians, that physicians may be unaware of their lack of knowledge and skills, or that physicians may have a lower demand for in-depth knowledge about PGx in general compared to pharmacists. Further research is needed to investigate the details underlying this barrier.

Literature reports that recognition of the clinical utility of PGx is a facilitator for implementation and that disbelief is a barrier.^8,9,10,12,13^ In our study, patients were positive about PGx, including its expected clinical utility, regardless of the occurrence of DGIs during their treatment, whereas HCPs were generally positive about the clinical utility, although some did not feel ready to apply it in daily practice. These results should, however, be interpreted with caution, because patients and HCPs who recognized the clinical utility were more likely to participate in this study and our study size was limited. In addition, patients and physicians were recruited from only two outpatient clinics, Psychiatry and Internal Medicine, and this may have influenced outcome, for example because practical use of reactive PGx testing is relatively common in psychiatry compared to other medical fields.

In our study, PGx screening results were reported directly to patients by mail without presence of an HCP. In the absence of a standardized reporting format for PGx testing results, which has previously been reported as a barrier^9^, we drafted a patient result letter with a brief explanation of the results in laymen’s terms and suggested actions, e.g. that the patient discuss their results with their current HCPs and share results with any new ones. Considering that pharmacotherapy is often a complex balance between treatment options, effectiveness, (risk of) ADRs, co-morbidities, and co-medication, it is our view that communicating the implications should be up to the individual HCP and should be tailored to the individual patient at the time it is relevant. Patients should only have to know when to share the PGx screening results with their HCP, e.g. in those cases where that information is not routinely included in their EHR. While the patient result letter was developed based on feedback from patients in focus groups prior to the study, our results indicate that some patients wanted to receive more and different information than provided. Most importantly, patients repeatedly reported wanting to know the exact implication of the PGx screening results for them, e.g. the level of dose adjustment or suitable alternative drugs. However, not all patients desired this depth of information, implying that one format for reporting PGx results to all patients would not suffice. An electronic personal health environment could present information to patients about their PGx screening results while containing multiple layers of information that enable them to receive the depth of information they desire, while also providing a standardized reporting format for PGx results and way for patients to easily share their results with their HCPs.

A new barrier emerged from our study: the unclear allocation of responsibilities among HCPs. The majority of patients reported that PGx screening results should be discussed with them by an HCP, but had differing preferences for which HCP should be responsible. We also found that HCPs themselves perceived they had a shared, and therefore still unclear, responsibility for discussing PGx screening results with patients. It was also unclear to both patients and HCPs at what point in the treatment process PGx screening results and their implications should be discussed, if ever. It is also unclear which HCP is ultimately responsible for the application of PGx screening results in different patient care situations. Furthermore, a group of patients reported their current drugs were not reviewed by an HCP even though they desired this (data presented in **Supplementary Results** section 1). Although some patient’s drugs may have been reviewed without their knowledge, these results underline the importance of clear communication with patients and expectation management. In addition, we should be aware of the risk of suboptimal pharmacotherapy in situations where patients are unassertive or have a more “wait-and-see” attitude, because it is unclear which HCP is responsible for discussing and applying PGx in practice. In our opinion, it should never be the patient’s responsibility to make sure PGx screening results are discussed and/or applied. Overall, this newly identified barrier needs to be addressed to facilitate responsible implementation of PGx screening. However, this may not be easily done nationally or internationally, as the interactions between HCPs can be highly variable between countries, regions, and even healthcare organizations or HCPs. As we identified this barrier in our limited local setting, additional research is needed to identify whether an unclear allocation of responsibilities is also a national/international barrier.

For logistical reasons, CDS software was only available as a separate tool outside the EHR in which the drugs are prescribed during our study, which presented a barrier for physicians to consider PGx screening results during prescription. This approach was taken because the availability of CDS software was deemed crucial in our pre-pilot study (see **Supplementary Methods** section 2), which is supported by literature.^7,12,13^ In response to our explorative pilot study, PGx-based medication surveillance has now been incorporated into our hospital EHR (since July 2020) in order to facilitate application of DPWG guidelines for every patient, both those admitted and those treated in outpatient clinics. The availability of CDS within our EHR is an important and crucial step towards use of PGx-based medication surveillance in routine healthcare. However, not all computer systems used by HCPs outside of our hospital can handle (all) PGx screening results. Since HCPs rely heavily on their computer system for insight into DPWG guidelines during drug prescription and medication surveillance, the lack of availability of CDS might be an important barrier within the Dutch healthcare in general.

In the Netherlands, PGx testing is currently only reimbursed by the insurer to investigate the cause of an ADR or as part of an optional reimbursement package. In anticipation of resolving this financial barrier to broad implementation of PGx testing and screening, we provided physicians with the opportunity to perform PGx screening for their patients free-of-charge and with minimal selection criteria. This study did not address which patients should be screened and at what timepoint in their treatment the costs of PGx screening would be best justified. Further research, including health technology assessment, should inform policy decision-making on these aspects.

To conclude, our exploratory pilot study confirmed known practical barriers and facilitators and suggested a new barrier to the implementation of PGx screening, namely an unclear allocation of responsibilities among HCPs. With this knowledge, we have more insight into which facilitators can be leveraged and which barriers need to be overcome to successfully implement PGx screening in Dutch outpatient hospital care. This study also provides a foundation for more detailed novel research that will hopefully further aid PGx implementation and contribute to unlocking the full potential of genome-guided drug prescription to enable personalized medication schemes with optimized treatment tolerance and response.

### STUDY HIGHLIGHTS

What is the current knowledge on the topic?

*There is limited practical experience with PGx in daily Dutch practice even though national guidelines on the practical interpretation of PGx results have been available since 2006*.

What question did this study address?

*What are the practical barriers and facilitating factors for HCPs and patients in the implementation of PGx screening in Dutch outpatient hospital care?*

What does this study add to our knowledge?

*This study provides insight into which facilitators can be leveraged and which barriers need to be overcome to successfully implement PGx screening in outpatient hospital care and provides a foundation for further, more detailed research into practical barriers and facilitators to implementation of PGx screening. For example, we observed that patients had a positive attitude toward PGx regardless of the occurrence of DGIs during their treatment*.

How might this change clinical pharmacology or translational science?

*Implementing PGx can unlock the full potential of genome-guided drug prescription, which will enable personalized medication schemes with optimized treatment tolerance and response*.

## Supporting information

Supplementary Results

## Data Availability

Data is available upon reasonable request.

## ACKNOWLEDGEMENTS

The authors thank Kate Mc Intyre for editing the manuscript, the UMCG Department of Genetics Integral Sample Management for processing DNA samples, and the UMCG Departments of Internal Medicine, Psychiatry and Clinical Pharmacy and Pharmacology, community pharmacists and patients for participating in this research. We thank the UMCG Genomics Coordination Center, the UG Center for Information Technology, and their sponsors BBMRI-NL & TarGet for storage and compute infrastructure.

The authors report no conflict of interest.

This research was supported by a University Medical Center Groningen (UMCG) Healthy Ageing Pilot fund (CDO15.0022) and by a Netherlands Organization for Scientific Research Spinoza Prize (SPI 92-266 to C.W.).

PL wrote the manuscript with critical input from all authors.

PL, RAS, ROBG, JGWK, MP, IMvL, AVR, BW and RHS designed the research.

PL, WHD and LGB performed the research.

PL, AvH, AMAH and AEP analyzed the data.

WHD and GvdV contributed new analytical tools.

MP, AVR, LHF, BW and RHS supervised data analysis.

## SUPPLEMENTARY MATERIALS

Supplementary Material 1: **Printed patient information (in Dutch)**

Supplementary Material 2: **Informed consent form (in Dutch)**

Supplementary Material 3: **Transcript of project website (in Dutch)**

Supplementary Material 4: **Animated video (in Dutch)**

Supplementary Material 5: **Patient result letter (English translation)**

Supplementary Methods

Supplementary Results

Table S1: **Details on custom genotyping panel**

Table S2: **Survey items** [Overview of the questions included in the surveys]

Table S3: **Demographics of study participants** [Demographics of participating patients, community pharmacists, hospital pharmacists and physicians]

Table S4: **Frequencies of PGx haplotypes and predicted phenotypes**

## REFERENCES

1. Bartlett, M. J., Green, D. W. & Shephard, E. A. Pharmacogenetic testing in the UK clinical setting. The Lancet 381, 1903 (2013).

2. Rigter, T. et al. Implementation of Pharmacogenetics in Primary Care: A Multi-Stakeholder Perspective. Front Genet 11, 10 (2020).

3. Krebs, K. & Milani, L. Translating pharmacogenomics into clinical decisions: do not let the perfect be the enemy of the good. Hum Genomics 13, 39 (2019).

4. Swen, J. J. et al. Pharmacogenetics: from bench to byte. Clin Pharmacol Ther 83, 781–7 (2008).

5. Swen, J. J. et al. Pharmacogenetics: from bench to byte--an update of guidelines. Clin Pharmacol Ther 89, 662–73 (2011).

6. Bank, P. C. D., Swen, J. J. & Guchelaar, H. J. Estimated nationwide impact of implementing a preemptive pharmacogenetic panel approach to guide drug prescribing in primary care in The Netherlands. BMC Med 17, 110 (2019).

7. Gelder, T. van & Schaik, R. H. N. van [Pharmacogenetics in daily practice]. Ned. Tijdschr. Geneeskd. 164, (2020).

8. Wouden, C.H. van der et al. Assessing the Implementation of Pharmacogenomic Panel-Testing in Primary Care in the Netherlands Utilizing a Theoretical Framework. J. Clin. Med. 9, 814 (2020).

9. Shuldiner, A. R. et al. The Pharmacogenomics Research Network Translational Pharmacogenetics Program: Overcoming Challenges of Real-World Implementation. Clin. Pharmacol. Ther. 94, 207–210 (2013).

10. Stanek, E. J. et al. Adoption of Pharmacogenomic Testing by US Physicians: Results of a Nationwide Survey. Clin. Pharmacol. Ther. 91, 450–458 (2012).

11. Van Driest, S. et al. Clinically Actionable Genotypes Among 10,000 Patients With Preemptive Pharmacogenomic Testing. Clin. Pharmacol. Ther. 95, 423–431 (2014).

12. Horgan, D. et al. An Index of Barriers for the Implementation of Personalised Medicine and Pharmacogenomics in Europe. Public Health Genomics 17, 287–298 (2014).

13. Dunnenberger, H. M. et al. Preemptive Clinical Pharmacogenetics Implementation: Current Programs in Five US Medical Centers. Annu. Rev. Pharmacol. Toxicol. 55, 89–106 (2015).

14. Ajzen, I. The theory of planned behavior. Organ. Behav. Hum. Decis. Process. 50, 179–211 (1991).

15. Sytema, S. & Krieke, L. van der Routine outcome monitoring: A tool to improve the quality of mental health care? In Improv. Ment. Health Care (Thornicroft, G., Ruggeri, M. & Goldberg, D.) 246–263 (John Wiley & Sons, Chichester, UK, 2013).doi:10.1002/9781118337981.ch16

16. R Core Team R: A Language and Environment for Statistical Computing. (R Foundation for Statistical Computing, Vienna, Austria, 2018).at <https://www.r-project.org/>

17. Sediq, R. et al. Concordance assessment of self-reported medication use in the Netherlands three-generation Lifelines Cohort study with the pharmacy database iaDB.nl: The PharmLines initiative. Clin Epidemiol 10, 981–989 (2018).

