## Supplementary Material 1 for "Practical barriers and facilitators experienced by patients, pharmacists and physicians to the implementation of pharmacogenomic screening in Dutch outpatient hospital care – an explorative pilot study"

### Doet u mee aan het wetenschappelijk onderzoek **MEDICATIE OP MAAT?**

*Op basis van uw DNA sneller het juiste medicijn en de juiste dosis bepalen*

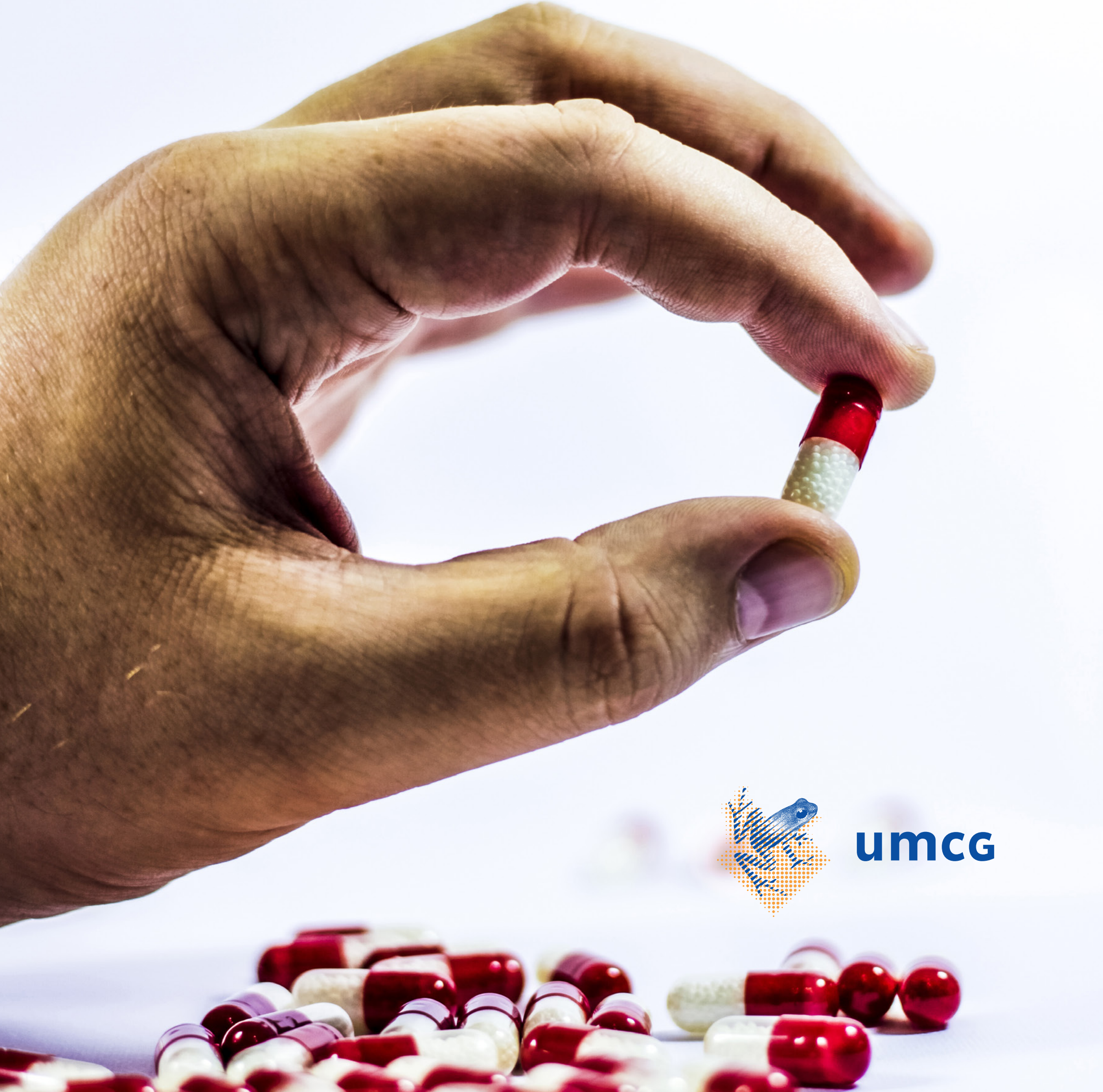

umcg

#### Vragen

Heeft u vragen of opmerkingen over het onderzoek?
Neem dan contact op met het team van Medicatie op Maat.

Mw. drs. Pauline Lanting, apotheker-onderzoeker

Telefoonnummer: (050) 361 07 06
Bereikbaar op maandag t/m donderdag van 9.00 - 17.00 uur.

#### www.medicatieopmaat.umcg.nl

Kijk voor meer informatie over het onderzoek op onze website. Hier staat ook een animatie waarmee Medicatie op Maat wordt uitgelegd.

#### Inleiding

U ontvangt deze folder omdat u kunt meedoen aan het wetenschappelijk onderzoek Medicatie op Maat. We beschrijven hier wat het onderzoek inhoudt, wat u ervan mag verwachten en hoe u zich hiervoor kunt aanmelden.

#### Wat onderzoeken we bij Medicatie op Maat?

Als u medicijnen krijgt voorgeschreven, houden we met verschillende dingen rekening. Denk hierbij aan uw leeftijd, geslacht, ziektebeeld en of u al andere medicijnen gebruikt.

Niet iedereen reageert altijd zoals verwacht op het voorgeschreven medicijn. Als het medicijn niet voldoende werkt of als u last heeft van bijwerkingen, kan de arts de dosis veranderen of andere medicijnen proberen. Hier gaat vaak wat tijd overheen.

Het doel van het onderzoek Medicatie op Maat is om per patiënt sneller het juiste medicijn en de juiste dosis te vinden. Door de relatie tussen uw erfelijk materiaal (DNA) en de reactie op medicatie te onderzoeken, kunnen we beter voorspellen hoe u gaat reageren op medicijnen. Met deze kennis kan uw medicijngebruik in de toekomst beter op uw persoonlijke situatie afgestemd worden.

#### Medicijnen in uw lichaam

Zodra u een medicijn inneemt, wordt het opgenomen in het bloed. Het bloed brengt een medicijn door uw hele lichaam. Uw stofwisseling zorgt er daarna voor dat het medicijn wordt afgebroken. Het volgende kan gebeuren:

- Het medicijn heeft voldoende tijd in uw lichaam om te werken en wordt vervolgens afgebroken.
- Het medicijn wordt te langzaam afgebroken waardoor het langer in het lichaam blijft dan nodig is. Het medicijn werkt vaak wel, maar u kunt last krijgen van bijwerkingen.
- Het medicijn wordt te snel afgebroken, waardoor het korter in het lichaam is dan noodzakelijk. Hierdoor werkt het medicijn minder goed of helemaal niet.

Per medicijn (of combinatie van medicijnen) kan de situatie per persoon verschillen. Om deze reden hebben sommige patiënten een andere dosis of een ander medicijn nodig om het gewenste effect te bereiken.

#### De rol van DNA

Uw erfelijk materiaal (DNA) bepaalt hoe uw stofwisseling verloopt. Dus uw DNA bepaalt mede hoe medicijnen in uw lichaam worden verwerkt. DNA zit onder andere in uw bloed. Door uw bloed te onderzoeken, wordt duidelijker hoe u kunt reageren op medicatie. Hierdoor kunnen we in de toekomst sneller het juiste medicijn en de juiste dosis voor u vinden.

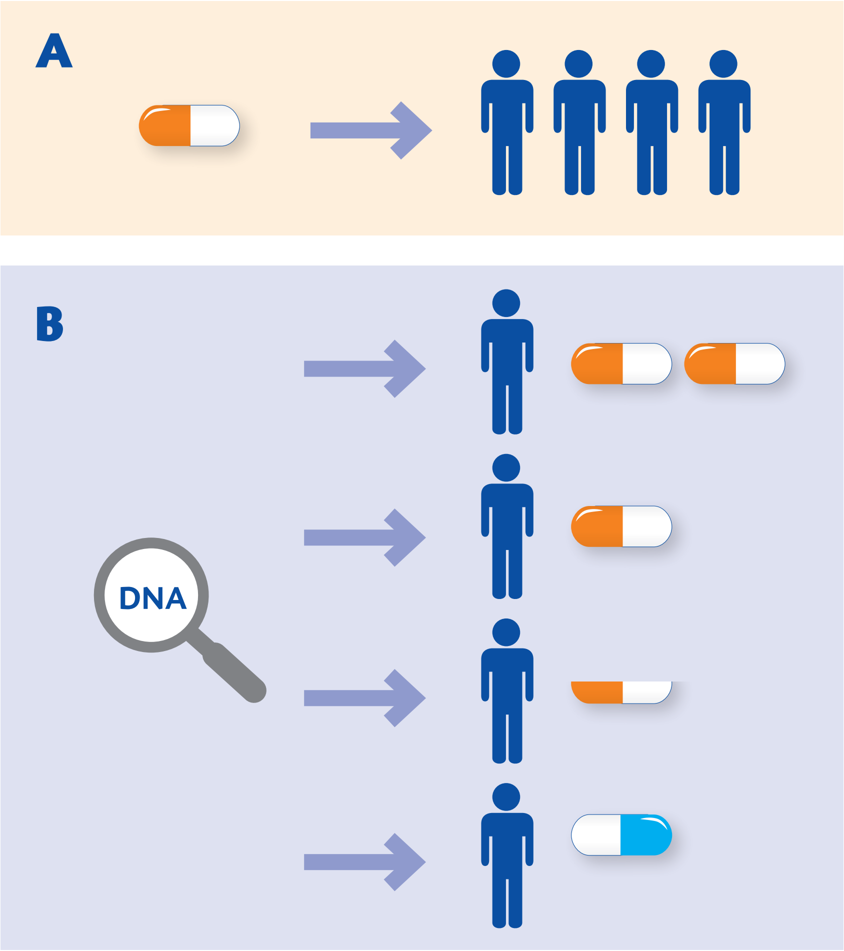


In bovenstaande figuur zien we twee situaties.
Bij situatie A gaat hetzelfde medicijn naar alle patiënten met dezelfde klachten. Niet bij iedereen wordt het gewenste effect bereikt. Bij situatie B is er gekeken naar het DNA van iedere patiënt. Het persoonlijke DNA laat zien dat sommige patiënten een andere dosis of een ander medicijn nodig hebben om het gewenste effect te bereiken. Hierdoor is er een kleinere kans op bijwerkingen en een grotere kans dat het medicijn goed werkt.

#### Meedoen aan het onderzoek

U kunt meedoen aan dit onderzoek als u 18 jaar of ouder bent en Nederlands spreekt. Als u wilt meedoen vult u het toestemmingsformulier in, dit zit bij de uitnodigingsbrief. Het toestemmingsformulier geeft u aan uw arts tijdens uw afspraak op de polikliniek.

#### Wat verwachten wij van u?

Als u meedoet gaat u na de afspraak gelijk naar de prikpoli om één buisje bloed te laten afnemen. Het bloed sturen we vervolgens op naar de afdeling Genetica in het UMCG, waar uw DNA onderzocht wordt. Omdat deze DNA-test voor het eerst in de praktijk wordt gebracht, zijn wij erg benieuwd naar uw mening over de DNA-test en de communicatie daarover. Daarom vragen wij u om twee vragenlijsten in te vullen. Uw reactie helpt ons om deze DNA-test in de toekomst nog verder te verbeteren en hopelijk voor meer patiënten beschikbaar te stellen.

#### Wat onderzoeken we in uw bloed?

Bij de DNA-test kijken we alleen naar variaties in uw DNA die van invloed zijn op de reactie op medicatie. Dat betekent dat er geen andere informatie uit uw DNA wordt onderzocht of waargenomen.

De variaties in het DNA die wij onderzoeken, beïnvloeden een beperkt aantal medicijnen. Dit kan betekenen dat het onderzoek niet direct invloed heeft op uw huidige medicatie. Het is mogelijk dat u in de toekomst medicijnen nodig heeft, waarvoor we door dit onderzoek beter kunnen voorspellen hoe u hierop reageert.

#### Uitslag van het onderzoek

Ongeveer zes weken na de DNA-test ontvangt u een brief met de uitslag. Uw behandelaars in het UMCG, uw huisarts en uw apotheek ontvangen deze gegevens ook. De uitslag kunt u zonodig bespreken met uw behandelaar in het UMCG, huisarts of apotheker.

De uitslag wordt opgeslagen in uw medisch dossier in het UMCG. Als u nieuwe medicatie nodig heeft, kan het computer-systeem een melding geven als u ergens over- of ongevoelig voor bent. Met die informatie kunnen uw behandelaars u dus medicatie op maat bieden.

#### Kosten

Er zijn voor u geen kosten verbonden aan de DNA-test, omdat deze test onderdeel is van wetenschappelijk onderzoek.

#### Privacy

De uitslag van de DNA-test zetten we in uw medisch dossier, dat alleen toegankelijk is voor uw behandelaars in het UMCG. Pas bij het voorschrijven van nieuwe medicatie worden deze gegevens gebruikt om uw medicijngebruik beter op uw persoonlijke situatie af te stemmen.

Overgebleven bloed of DNA wordt na het onderzoek vernietigd.

Universitair Medisch Centrum Groningen
