## Supplementary Material 2 for "Practical barriers and facilitators experienced by patients, pharmacists and physicians to the implementation of pharmacogenomic screening in Dutch outpatient hospital care – an explorative pilot study"

**Toestemmingsformulier voor de deelnemer aan het onderzoek**

### Medicatie op Maat

- Ik heb de informatiebrief gelezen. Ook kon ik vragen stellen. Mijn vragen zijn voldoende beantwoord. Ik had genoeg tijd om te beslissen of ik meedoe.
- Ik weet dat meedoen vrijwillig is. Ook weet ik dat ik op ieder moment kan beslissen om toch niet mee te doen of te stoppen met het onderzoek. Daarvoor hoef ik geen reden te geven.
- Ik geef toestemming om mijn huisarts/specialist(en) en apotheker te informeren over de vastgestelde erfelijke eigenschappen met betrekking op mijn medicatie.
- Ik weet dat sommige mensen mijn gegevens kunnen inzien. Die mensen staan vermeld in de informatiebrief.
- Ik geef toestemming voor het verzamelen en gebruiken van mijn gegevens en bloedmonster op de manier en voor de doelen die in de informatiebrief staan.
- Ik wil meedoen aan dit onderzoek.
- **Ik wil wel/niet* in de toekomst benaderd worden voor eventueel vervolgonderzoek**
  ***doorhalen wat niet van toepassing is**

**Naam deelnemer:** ________________________________________

**UMCG-nummer deelnemer:** ______________________

**Handtekening deelnemer:** **Datum:** __ / __ / __

------------------------------------------------------------------------------------------------------------

*Het onderstaande deel wordt ingevuld door het onderzoeksteam*

Ik verklaar dat ik deze deelnemer volledig heb geïnformeerd over het genoemde onderzoek.

Als er tijdens het onderzoek informatie bekend wordt die de toestemming van de deelnemer zou kunnen beïnvloeden, dan breng ik hem/haar daarvan tijdig op de hoogte.

Naam onderzoeker (of diens vertegenwoordiger):

Handtekening onderzoeker (of diens vertegenwoordiger): Datum: __ / __ / __

------------------------------------------------------------------------------------------------------------

*De deelnemer krijgt een kopie van het getekende toestemmingsformulier thuisgestuurd.*
