## Supplementary Material 3 for "Practical barriers and facilitators experienced by patients, pharmacists and physicians to the implementation of pharmacogenomic screening in Dutch outpatient hospital care – an explorative pilot study"

umcg Genetica

### Medicatie op Maat

Lees voo

Animatie Medicatie op Maat

Bij Medicatie op Maat onderzoekt het UMCG de relatie tussen iemands erfelijk materiaal (DNA) en zijn reactie op medicatie. Door bloed te onderzoeken wordt duidelijker hoe iemand kan reageren op medicatie. Het doel van Medicatie op Maat is om per patiënt sneller het juiste medicijn en de juiste dosis te vinden.

#### Waarom DNA-onderzoek?

Bij dit wetenschappelijk onderzoek doen we een DNA-test. Dit houdt in dat we op zoek gaan naar variaties in het DNA. Deze variaties kunnen verklaren waarom bepaalde medicijnen bij sommige mensen niet of niet goed werken. Ook kunnen deze verklaren waarom sommige mensen bijwerkingen krijgen. Het persoonlijke DNA laat zien dat sommige patiënten een andere dosis of een ander medicijn nodig hebben om het gewenste effect te bereiken. Hieronder vindt u een filmpje waarin Medicatie op Maat wordt uitgelegd.

#### Wie kunnen meedoen?

U kunt alleen op uitnodiging deelnemen aan dit onderzoek. We vragen patiënten van de afdelingen Psychiatrie, Ouderengeneeskunde of Interne Geneeskunde van het UMCG. Ook als u geen medicijnen gebruikt kunt u gevraagd worden mee te doen aan Medicatie op Maat. Mogelijk krijgt u in de toekomst een medicijn voorgeschreven.

#### Wat levert deelname u op?

Als u instemt met deelname, dan weet u na de test welke medicijnen naar verwachting het beste bij u passen. Hierdoor is er een kleinere kans op bijwerkingen en een grotere kans dat het medicijn goed werkt.
Bij deze DNA-test kijken we alleen naar variaties in het DNA die van invloed zijn op de reactie op medicatie. Dat betekent dat er geen andere informatie uit het DNA wordt onderzocht of waargenomen.

---

Deel dit:

### Lijst van medicijnen

Hieronder vindt u een overzicht van de medicijnen waarop een invloed van genen bekend is. Doordat er (wereldwijd) veel onderzoek gedaan wordt naar de invloed van genen op medicijnen wordt deze lijst steeds langer.

Lees vo

#### Overzicht medicijnen waarvan invloed genen bekend is

### A

- Abacavir
- Acenocoumarol
- Amitriptyline
- Anticonceptie met oestrogenen
- Aripiprazol
- Atomoxetine
- Atorvastatine
- Azathioprine / Mercaptopurine

### C

### D

### E

### F

### G

### H

### I

### L

### M

### N

### O

### P

### Q

### R

### S

### T

### V

### W

### Z

Deel dit:

### Informatie voor zorgverleners

Lees vo

Het onderzoek Medicatie op Maat is een implementatie studie van farmacogenetische screening in de klinische praktijk van het UMCG. Het betreft een pilot onderzoek van een jaar met een beperkte omvang. Vooralsnog kunnen alleen patiënten van de afdelingen Psychiatrie, Ouderengeneeskunde en Interne Geneeskunde van het UMCG deelnemen. Indien uw patiënt niet tot deze afdelingen behoort en u wel geïnteresseerd bent in een farmacogenetische screening, dan is dit mogelijk via de reguliere weg.

#### Toepassen van de DNA-uitslag

- **Voorschrijven volgens de farmacogenetica richtlijn**
  Om toepassing van de uitslagen in het UMCG op het moment van voorschrijven te vereenvoudigen is Clinical Decision Support software ontwikkeld. Na invoer van een geneesmiddel zal op basis van de DNA-uitslag en de G-standaard al dan niet een advies getoond worden. De getoonde adviezen zijn onderdeel van de G-standaard en opgesteld door de KNMP werkgroep farmacogenetica.
- **Medicatiebewaking**
  De medicatiebewaking van poliklinische voorschriften valt nadrukkelijk onder de verantwoordelijkheid van de openbaar apotheker. Indien met een voorschrift wordt afgeweken van de gebruikelijke werkwijze, dient op het recept te worden vermeld dat rekening is gehouden met het farmacogenetisch profiel van de patiënt.

#### Contact

Voor advies betreffende de behandeling van een patiënt kan contact worden opgenomen met de ziekenhuisapotheek (apotheker farmaceutische patiëntenzorg: 55751). Voor overige vragen kunt u contact opnemen met het team van Medicatie op Maat (, tel. projectcoördinator: 10706).

#### Genpanel en relevante medicatie

De DNA-test bestaat uit een panel van 15 genen (zie onderstaande tabel), overeenkomend met de KNMP richtlijn farmacogenetica. Bij meer dan 60 medicijnen is sprake van een zogenaamde gen-geneesmiddel-interactie (GGI) en is farmacogenetica relevant. In de richtlijn zijn ook een aantal medicijnen opgenomen waarvoor geen sprake is van een GGI, hiervoor is vastgesteld dat farmacogenetica niet relevant is. Een volledig overzicht van de richtlijn is te raadplegen via de KNMP kennisbank .

**CYP1A2**

**CYP2B6**

**CYP2C9**

**CYP2C19**

**CYP2D6**

**CYP3A4**

**CYP3A5**

**DPYD**

**Fact. V**

**HLA**

**MTHFR**

SLCO1B1

TPMT

UGT1A1

VKORC1

Deel dit:

### Betrokken onderzoeken

Lees vo

#### Afdelingen

Het onderzoek Medicatie op Maat is een samenwerking tussen de afdelingen Genetica, Klinische Farmacie en Farmacologie, Psychiatrie, Ouderengeneeskunde en Interne Geneeskunde van het UMCG.

De afdeling Genetica voert de DNA-test uit en voert het wetenschappelijk onderzoek uit dat hoort bij Medicatie op Maat. Ook wordt het onderzoek gecoördineerd en ondersteund vanuit deze afdeling.

De afdeling Klinische Farmacie en Farmacologie is binnen het ziekenhuis verantwoordelijk voor allerlei zaken rondom medicatie. De apotheek van het ziekenhuis hoort bij deze afdeling en hier werken dan ook veel (ziekenhuis)apothekers die artsen kunnen adviseren over Medicatie op Maat. De (ziekenhuis)apothekers zijn deelnemers van het wetenschappelijk onderzoek en zullen gaan werken met de uitslagen van de DNA-test. Een vertegenwoordiger van deze afdeling is als deskundige betrokken bij de uitvoering van het wetenschappelijk onderzoek, maar is zelf geen deelnemer van het onderzoek.

De afdelingen Psychiatrie, Ouderengeneeskunde en Interne Geneeskunde zijn deelnemers van het wetenschappelijk onderzoek dat hoort bij Medicatie op Maat. Zij zullen als eerste afdelingen in het ziekenhuis gaan werken met de uitslagen van de DNA-test.

#### Het onderzoeksteam

Het onderzoek Medicatie op Maat wordt uitgevoerd door een team van deskundigen werkzaam bij de afdelingen Genetica en Klinische Farmacie en Farmacologie.

---

**Deel dit:**
