## Supplementary Material 5 for "Practical barriers and facilitators experienced by patients, pharmacists and physicians to the implementation of pharmacogenomic screening in Dutch outpatient hospital care – an explorative pilot study"

**Results 'Personalized Medicine'**

Datum: 11 november 2020

Dear [name],

Some time ago you had a blood sample taken for the scientific research study 'Personalized Medicine'. We have now tested your blood in the laboratory of the Genetics Department of the UMCG, and this letter explains the results of the test.

**About the test**

We tested 14 genes that may be of importance for the use of certain drugs. This makes it possible to better predict how you will respond to these drugs.

In the appendix you will find an overview of the results of the test, which is intended for your healthcare provider(s). We keep this result in your medical file so that your healthcare practitioners at the UMCG can take this information into account from now on. We have also sent your results to the general practitioner and pharmacy known to us. You can also show this result to your other healthcare providers, such as doctors in other hospitals or the thrombosis service. They can use it to adjust your medication to your hereditary sensitivity.

**Your result**

The tables on the next page list the medicines for which we can currently predict your hereditary susceptibility. In the future, the number of medicines for which we can do this will continue to increase. The list below is therefore a snapshot of what we know at present. If you are already taking (one of) these medicines, this information does not mean that anything needs to be changed. It may be that the dosage has already been adjusted sufficiently to your personal situation.

The result only tells you whether you have a normal or abnormal hereditary sensitivity to certain medicines. You may be more or less sensitive, depending on the drug. Your doctor and pharmacist can tell you more about this. Using the enclosed results, your medicines can be adjusted to your hereditary sensitivity, where necessary. Your doctor and pharmacist will also directly consider other factors that play a role in your response to medication, such as your kidney function, illnesses you have, or other medications you are taking.

*Important! Never adjust your medication yourself. If you are unsure about the dosage or drug, always discuss this with your doctor or pharmacist.*

| You have a normal hereditary sensitivity to the following medicines: | |
|---|---|
| Abacavir | Antiviral medication |
| Acenocoumarol | Blood thinner |
| Amitriptyline | Antidepressant |
| Contraceptive pill with estrogen | 'the pill' |
| Aripiprazole | Antipsychotic |
| Atomoxetine | ADHD medication |
| Atorvastatin | Cholesterol lowering medication |
| Azathioprine | Immune suppressant |
| Citalopram | Antidepressant |
| Clomipramine | Antidepressant |
| Clopidogrel | Blood thinner |
| Codein | Painkiller |
| Doxepin | Antidepressant |
| Efavirenz | Antiretroviral medication |
| Eliglustat | Medication for metabolic disease |
| Escitalopram | Antidepressant |
| Phenprocoumon | Blood thinner |
| Phenytoine | Antiepileptic |
| Flecainide | Cardiac arrythmia medication |
| Flucloxacillin | Antibiotic |
| Haloperidol | Antipsychotic |
| Imipramine | Antidepressant |
| Irinotecan | Cancer medication |
| Lansoprazole | Antacid |
| Mercaptopurine | Cancer medication and immune suppressant |
| Metoprolol | High blood pressure medication |
| Nortriptyline | Antidepressant |
| Omeprazole | Antacid |
| Oxycodone | Painkiller |
| Pantoprazole | Antacid |
| Paroxetine | Antidepressant |
| Pimozide | Antipsychotic |
| Propafenon | Cardiac arrythmia medication |
| Sertraline | Antidepressant |
| Simvastatin | Cholesterol lowering medication |
| Tacrolimus | Immune suppressive medication |
| Tamoxifen | Cancer medication |
| Tramadol | Painkiller |
| Venlafaxine | Antidepressant |
| Voriconazole | Antifungal |
| Warfarin | Blood thinner |
| Zuclopentixol | Antipsychotic |

| You have an abnormal hereditary sensitivity to the following medicines | |
|---|---|
| Abacavir | Antiviral |

| | |
|---|---|
| Acenocoumarol | Blood thinner |
| Amitriptyline | Antidepressant |
| Contraceptive pill with estrogen | 'The Pill' |
| Aripiprazole | Antipsychotic |
| Atomoxetine | ADHD medication |
| Atorvastatin | Cholesterol lowering medication |
| Azathioprine | Immune suppressive |
| Citalopram | Antidepressant |
| Clomipramine | Antidepressant |
| Clopidogrel | Blood thinner |
| Codeine | Painkiller |
| Doxepin | Antidepressant |
| Efavirenz | Antiviral |
| Eliglustat | Medication for metabolic disease |
| Escitalopram | Antidepressant |
| Phenprocoumon | Blood thinner |
| Phenytoine | Antiepileptic |
| Flecainide | Cardiac arrythmia medication |
| Flucloxacillin | Antibiotic |
| Haloperidol | Antipsychotic |
| Imipramine | Antidepressant |
| Irinotecan | Cancer medication |
| Lansoprazole | Antacid |
| Mercaptopurine | Cancer medication and immune suppressant |
| Metoprolol | Blood pressure lowering medication |
| Nortriptyline | Antidepressant |
| Omeprazole | Antacid |
| Oxycodone | Painkiller |
| Pantoprazole | Antacid |
| Paroxetine | Antidepressant |
| Pimozide | Antipsychotic |
| Propafenon | Cardiac arrythmia medication |
| Sertraline | Antidepressant |
| Simvastatin | Cholesterol lowering medication |
| Tacrolimus | Immune suppressant |
| Tamoxifen | Cancer medication |
| Tramadol | Painkiller |
| Venlafaxine | Antidepressant |
| Voriconazole | Antifungal |
| Warfarin | Blood thinner |
| Zuclopentixol | Antipsychotic |

**Questions?**

If you have any questions about the results, you can discuss them with your treating doctor at the UMCG. You may also contact the researchers of the study. For this you can send an e-mail to [e-mail address] or call [phone number].

Kind regards on behalf of the Customized Medication team,

Prof. R.H. Sijmons, Clinical Geneticist
Professor of Medical Translational Genetics UMCG

Prof. B. Wilffert, clinical pharmacologist
Professor of Pharmacotherapy and Clinical Pharmacy UMCG

*Appendix: Result of pharmacogenetic screening (information for healthcare providers)*

### Appendix: Results pharmacogenetic screening

#### Information for healthcare providers

*Origin of this result*
This result has been provided as part of the scientific research study ‘Personalized Medicine’. This is a pilot study into outpatient implementation of pharmacogenetic screening in the UMCG. The genetic variants included in this screening are based on the DPWG guidelines (reference date: December 2016). If you still have questions or uncertainties about your patient’s pharmacotherapy after consulting the guidelines and consultation between the patient’s doctor and pharmacy, the hospital pharmacy of the UMCG is available to give advice to colleagues. For this, you can contact us by telephone ([phone number]). For all other questions or comments where you do not need advice about the treatment of your patient, please contact the project coordinator of the study ([e-mail address] or [phone number]). More information about the study is available via the project website [URL].

*Theoretical background*
Genetic variation in enzymes or drug targets can cause patients to respond differently to the same medication. For many enzymes, the genotype is translated into a predicted phenotype, the expected metabolic status. The term extensive metabolizer (EM) indicates a "normal" metabolism. A poor metabolizer (PM) has hardly any enzyme activity, an intermediate metabolizer (IM) has reduced enzyme activity and an ultra-rapid metabolizer (UM) has increased enzyme activity.

*Things to keep in mind about the result*
If the Dutch pharmacogenetic working group decides in the future to adjust the method of translation from genotype to phenotype, the phenotype in this result must be predicted again. Partly for this reason, it is also important to record the genotypes in medical records.

*Medication monitoring*
The guidelines for the application of pharmacogenetics are available via the KNMP Knowledge Base (Medication Monitoring - Pharmacogenetics) and included in the G-standard as contraindication. The results of this pharmacogenetic screening can be entered into digital information systems as contraindications. See www.knmp.nl/farmacogenetica for more information.

#### Result pharmacogenetic screening ‘Personalized Medicine’

Date: 11 november 2020

Name: UMCG-nr:

Birthdate:

| Gene | Genotype[1] | Predicted phenotype[2] | Alternative notation[3] |
|---|---|---|---|
| **CYP1A2**<br>Tested for *1C, *1F, *1L | | | - |
| **CYP2B6**<br>Tested for *6 | | | - |
| **CYP2C9**<br>Tested for *2, *3 | | | - |
| **CYP2C19**<br>Tested for *2, *3, *4A, *4B, *17 | | | - |
| **CYP2D6**[4]<br>Tested for *3, *4, *4M, *5, *6, *7, *9, *10, *12, *29, *36, *41, *69, *109 | | | - |
| **CYP3A4**<br>Tested for *22 | | | - |
| **CYP3A5**<br>Tested for *3, *6 | | - | |
| **Factor 5**<br>Tested for Leiden mutation | | - | |
| **HLA**<br>Tested for *5701 | | - | |
| **MTHFR**<br>Tested for 677C>T | | - | |
| **SLCO1B1**<br>Tested for 521T>C | | - | |
| **TPMT**[5]<br>Tested for *2, *3A, *3B, *3C | | | - |
| **UGT1A1**<br>Tested for *28, *36, *37 | | | - |
| **VKORC1**[6]<br>Tested for *2 | | - | |

[1] Determined following internationally applicable standards (Translation tables Clinical Pharmacogenetics Implementation Consortium).

[2] Determined following the current nationally applicable guidelines of the Dutch pharmacogenetics working group.

[3] If the G-standard uses a different notation than that reported under genotype or phenotype, it is shown here.

[4] CYP2D6: Based on the PCR technique used, it is not possible to distinguish between *10, *14, *37, *47, *49, *52, *54, *56B, *57, *65, *72, *87, *94, *95, *100, *101. The result is reported as *10 because the chances of this are higher. Metabolism other than that reported, due to rarer DNA variants, cannot be completely ruled out.

Based on the PCR technique used, it is not possible to distinguish between *41, *91. The result is reported as *41 because the chances of this are higher. Metabolism other than that reported, due to rarer DNA variants, cannot be completely ruled out.

[5] TPMT: Based on the PCR technique used, it is not possible to distinguish between *1/*3A (intermediate) and *3B/*3C (slow metabolism). The result is reported as *1/*3A because the chances of this are approximately 12.000x higher. Slow metabolism, due to rarer DNA variants, cannot be completely ruled out.

[6] VKORC1: Alternative notation of variants tested: -1639G>A. Medication monitoring is performed based on the variation at position 1173C>T that is inherited together with the tested variant. 1/*1 matches CC, *1/*2 matches CT, *2/*2 matches TT.

[7] We could not distinguish between different variants in the DNA. The variants that we could not distinguish do lead to the same predicted phenotype, so only the result of the genotype is inconclusive.
