## Supplementary Methods for "Practical barriers and facilitators experienced by patients, pharmacists and physicians to the implementation of pharmacogenomic screening in Dutch outpatient hospital care – an explorative pilot study"

1. *Study setting*

In the Netherlands, patients are commonly registered with one general practitioner (GP) and one community pharmacy. More specialized care is provided by medical specialists in outpatient clinics of general or academic hospitals. Prescriptions from physicians in the outpatient clinic are dispensed by community pharmacies. Patients can choose to collect their prescription at the community pharmacy where they are registered with or at any other community pharmacy as a visiting guest. With permission of the patient, pharmacies typically notify the pharmacy where a patient is registered of the drug(s) they have dispensed for visiting guests in order to maintain medication safety. In outpatient care, the GP typically receives a report from the hospital physician in which details are shared about the (differential) diagnosis, treatment plan, medication use, and relevant diagnostic measurements (i.e. blood pressure, drug plasma concentration, PGx test results).

2. *Qualitative interview and focus group studies*

An explorative qualitative interview and focus group study with 13 prescribers (from the participating outpatient clinics), 13 patients, and 7 pharmacists, preceded the development of the survey and final design of the implementation pilot study. In this pre-pilot study, we assessed barriers and facilitators as well as needs of prescribers, patients, and pharmacists, in the implementation of PGx.

A positive attitude towards clinical implementation of PGx was found among both professionals and patients. A lack in PGx knowledge among healthcare professionals was considered a barrier. Among pharmacists, despite their familiarity with the concept of PGx, there was lack of knowledge to confidently apply it in practice. For the prescribers, the availability of Clinical Decision Support (CDS) was regarded a critical success factor for implementing PGx in clinical practice. The distribution of responsibilities among professionals also needs addressing. Prescribers and pharmacists differed in views, both holding the other responsible. Also, there was a need among healthcare practitioners for an informative introductory meeting prior to the start of the study. In order to inform patients about PGx, an information brochure, animated video, and project website were developed and pilot-tested with patients. The offer of an animated video in addition to an information brochure and project website aided in understanding of PGx. Moreover, since the term PGx was found to be difficult, it was suggested we speak of personalized medication (in Dutch: Medicatie op Maat) in the implementation process.

3. *Recruitment of participants*

Physicians from the outpatient clinics of Internal Medicine and Psychiatry were asked to participate in this study and to recruit among their own patients. We choose these two outpatient clinics because they treat large volumes of patients, prescribe a substantial number of relevant drugs, and were enthusiastic about PGx implementation. Patients were recruited on a first-come-first-served basis until the maximum laboratory capacity of 165 was reached. The number of physicians was limited to 30 to increase the opportunity for participating physicians to include multiple patients and gain sufficient exposure to PGx. For practical reasons, genotyping was performed batch-wise, resulting in patients receiving their PGx screening results over the course of multiple months. Follow-up started from the time results were reported and lasted up to a year.

Inclusion was possible for adult patients proficient in Dutch, and patients were excluded when their physician predicted they would be overburdened by study participation or due to the inability to provide a blood sample. Note that no criteria were set regarding drug use or diagnosis.

Besides the patient, relevant stakeholders in outpatient care are the treating physician in the hospital and the community pharmacist because of their treatment relationship with the patient. Although the hospital pharmacy lacks official treatment relationship according to the Medical Treatment Agreement Act (WGBO), hospital pharmacists are available to consult with in-house physicians in outpatient clinics. For this reason, we decided to include the hospital pharmacist in our study as a relevant stakeholder. Although GPs have an official treatment relationship with the patients in our study, this applies to the care provided in primary care, and not to outpatient clinics. For this reason, we decided not to include the GP in our study as a relevant stakeholder.

The printed information and informed consent form that eligible patients received, transcripts of the project website, and the animated video are provided in the **Supplementary Materials.**

4. *Genotyping and reporting of test results*

DNA was isolated using the ReliaPrep™ Large Volume HT gDNA Isolation System with the Hamilton Microlab Star Plus instrument and was purified with isopropanol/ethanol if necessary. Single Nucleotide Polymorphism (SNP) genotyping was performed using a custom designed multiplex (iPLEX^®^) panel on the MassARRAY^®^ system (Agena Bioscience, San Diego, USA). Results were translated into PGx haplotypes and exported using PGxReport v2 with a custom translation table following Cytochrome P450 (CYP) allele nomenclature. Copy Number Variant (CNV) genotyping was performed for *CYP2D6* using pre-tested TaqMan^®^ Copy Number assays Hs04083572_cn, Hs04502391_cn, and Hs00010001_cn on the qPCR QuantStudio™ 7 Flex Instrument with QuantStudio™ software v1.2. Relative quantification of *CYP2D6* gene copy number was performed following the comparative C_T_ (ΔΔC_T_) method using CopyCaller v2.1. Variable Number Tandem Repeat (VNTR) genotyping for *UGT1A1* was performed using fragment length analysis with the Applied Biosystems Instrument 3730xl Genetic Analyzer.

SNP, CNV and VNTR genotyping results were integrated using a custom translation tool and registered in GLIMS (CliniSys, Chertsey, U.K.), the hospital’s laboratory information system. Subsequently, the PGx haplotypes were uploaded into the custom Clinical Decision Support software PGxConsultor which translated the haplotypes into predicted phenotypes based on translation tables following DPWG guideline background information. Both PGx haplotypes and predicted phenotypes were stored in the PGxConsultor database for use.

Patients received a letter explaining the PGx screening results in lay man’s terms. The result letter included explanation about the test performed, which drugs are (not) affected by the screening results, with whom the results are shared and where they are stored. PGx screening results were not reported to the patient on the level of the gene (i.e. PGx haplotype and predicted phenotype), but on the level of the drugs affected. We did include the actual results as an attachment so patients could share their results with other healthcare practitioners. This attachment meant for healthcare practitioners also included background information about the origin of the results, theoretical background and reference to the DPWG guidelines. A copy of the letter was made available in the electronic health record (EHR) for UMCG healthcare practitioners to use and send to their community pharmacist and GP. As a service to the participating physicians, they received an email when the result letter of their patient(s) became available in the EHR.

*5. Clinical Decision Support system*

PGxConsultor (Medecs BV, Eindhoven, the Netherlands) was developed as a stand-alone CDS system to provide prescribers with relevant Dutch Pharmacogenetics Working Group (DPWG) guideline recommendations for their patient, and uses the G-standard, the Dutch drug database which is used by all parties in healthcare in the Netherlands. The G-standard contains all products that are dispensed by or used in the pharmacy, as well as protocols for medication prescription including DPWG guidelines. The content of the G-standard is maintained and updated on a monthly basis by Z-index. PGxConsultor automatically updates to the latest version of the G-standard.

The user interface (UI) contains two input fields, one for the patient identifier and one for the drug of interest. For every search, the internal database with PGx haplotypes and predicted phenotypes of the patient of interest are compared with the G-standard. Whenever a DPWG guideline recommendation is found, this is displayed in the UI. In case of normal metabolizers, recommendation texts are absent and a default message is shown. This is also the case when the genotyping result was inconclusive. Together with the recommendation or default message, an optional message specific to our hospital is shown, for example information how to request laboratory tests if the recommendation includes that blood levels to be monitored. Finally, prescribers are asked to provide feedback on how they handled the information provided, for example whether they (partly) adhered to the recommendation. Every search was automatically registered in the study database for evaluation.

5. *Data collection*

Drug use during follow-up was extracted from the EHRs. Physicians use the EHR to prescribe drugs and are also able to record other drugs used at that time, for example drugs prescribed by the GP. While these drug records are supposed to be kept up-to-date, this is generally not the case and the most complete records can be found in the clinical notes and correspondence send out to the GP.
