## Supplementary Results for "Practical barriers and facilitators experienced by patients, pharmacists and physicians to the implementation of pharmacogenomic screening in Dutch outpatient hospital care – an explorative pilot study"

1. *Review of patient drug use in response to PGx screening results*

The survey study showed that 89% of patients reported using drugs at the time they received their PGx screening results, and 33% reported that their drugs were reviewed by a health care practitioner (HCP) in response to the results. No review was reported by 67% of patients, although 42% wanted their drugs reviewed. Patient reported reasons for lack of review were that they had not (yet) discussed their wish with their HCP(s) (53%), it was apparently not necessary (20%), drugs were changed anyway (7%), and unknown (20%). Initiative to review drugs was taken by either the physician (44%), patient (31%), community pharmacist (9%), or attendant caregivers (16%). Reviews were reported to have been performed by the hospital physician (71%), community pharmacist (16%), general practitioner (GP) (6%), a physician from another hospital (3%), or an unspecified HCP (3%). As a result of these reviews, 14 changes in drug treatment were reported by 36% (n=12) of the patients whose drugs were reviewed. Changes included switching to another drug (n=8), discontinuation of a drug (n=3), dose adjustments (n=2), and unspecified change (n=1). Eighteen patients (55%) whose drugs were reviewed but not changed reported the reason: no changes were needed (78%), risk of ADRs was unacceptable (6%), insufficient information to make a change (6%), or unspecified (11%). The remaining 9% of patients whose drugs were reviewed did not report any changes in drug treatment or why no changes were made.

2. *Clinical Decision Support searches and output*

Details of drug-gene interactions (DGIs) involved in clinical decision support (CDS) searches and resulting actions taken by physicians are presented in the table below.

One physician prescribed no alternative despite the recommendation to do so (marked with * in the table). (S)he commented that the indication for the drug was different from the indication for which the recommendation was written, there were no adverse drug reactions (ADRs), and the treatment was effective. If this would change, an alternative would be prescribed. This comment illustrates how the practical application of PGx can transcend guideline recommendations and application is not always straightforward.

Another physician prescribed an alternative medication to a patient already using a drug, in adherence with the recommendation shown (marked with # in the table). Looking into this particular case clearly shows the potential of PGx-guided drug prescription on a patient level. In this case, a 38-year old patient was diagnosed with small fiber peripheral neuropathy and treated with amitriptyline 30 mg. Therapy was ineffective and multiple ADRs were present. Genotyping revealed this patient as a CYP2D6 intermediate metabolizer, for which DPWG guidelines recommend prescribing an alternative if possible. After three weeks of pregabalin, the patient was pain-free and the ADRs were gone. The patient reported being ‘really happy’ to have participated in this study.

Amitriptyline is metabolized by CYP2C19 into the active metabolite nortriptyline. Both amitriptyline and nortriptyline are metabolized by CYP2D6. A lower metabolic capacity of CYP2D6 causes an increase in plasma concentrations of amitriptyline and nortriptyline and a decrease in plasma concentrations of the less potent or inactive metabolites.^1^

**Table SR1** Details of actionable recommendations

| Recommendation | DGIs involved | Resulting actions |
| --- | --- | --- |
| Adhere to adjusted maximum (daily) dose or prescribe alternative (n=6) | Citalopram + CYP2C19 (n=5)  Escitalopram + CYP2C19 (n=3)  Imipramine CYP2D6 (n=1) | No drug prescribed (n=3)  Alternative drug prescribed (n=1)  Drug stopped (n=1)  Drug already in use in adjusted maximum dose (n=1) |
| Prescribe alternative (n=5) | Venlafaxine + CYP2D6 (n=3)  Amitriptyline + CYP2D6 (n=2) | No drug prescribed (n=2)  Alternative drug prescribed (n=1)  Patient already using drug and switched to an alternative (n=1)#  No change made in patient already using drug (n=1)* |
| Lower dose and monitor plasma concentrations (n=4) | Nortriptyline + CYP2D6 (n=2) Imipramine + CYP2D6 (n=1) Clomipramine + CYP2D6 (n=1) | Blood drawn to determine plasma concentrations (n=2)  Drug prescribed (n=1)  Lower dose achieved via therapeutic drug monitoring in patient already using drug (n=1) |
| Adjust dose based on effect observed, increase dose, or lower maintenance dose (n=5) | Sertraline + CYP2C19 (n=2)  Phenytoin + CYP2C9 (n=1)  Omeprazole + CYP2C19 (n=1)  Metoprolol + CYP2D6 (n=1) | Drug not prescribed in any of these cases |

3. *Evaluation of DPWG guidelines*

HCPs found that the Dutch Pharmacogenetics Working Group (DPWG) guidelines were understandable (90%) and provided sufficient guidance to start treatment (86%), but insufficient guidance to continue treatment (75%). HCPs did not find it hard to decide what to do based on the recommendation shown when combined with their own knowledge (78%). PGx was not often an important clinical factor in pharmacotherapy (50%) and determining its place relevant to other clinical factors (i.e. kidney function) was not hard (60%). Overall, HCPs reported that DPWG guidelines can be implemented in their current form (70%).

4. *Practical application of PGx*

A patient was sent away by a pharmacy technician when showing their PGx screening results because (s)he ‘had never seen that before and did not know what to do with it’. The researchers were informed about this and contacted the pharmacy. The pharmacist registered the PGx screening results into the computer system and tested the warning signal. Coincidentally, right after this, a prescription with a DGI for this patient was processed by a (different) pharmacy technician. Since there was no mention of the DGI warning signal on the prescription, as local protocol dictates, the pharmacist consulted the pharmacy technician to check whether the warning was seen. The pharmacy technician had seen the warning but decided not to mention or act upon it since ‘(s)he had never seen a warning like that before and therefore did not think it was relevant’. This event clearly shows the importance of educating and informing all HCPs involved so that PGx warning signals are not ignored.

5. *Evaluation of the result letter*

After reading the result letter, 69% of patients reported understanding when to show the results to their HCPs, whereas 16% did not. 26% of patients reported doubting whether their current drugs are suitable for them, and 4% reported feeling less healthy. 52% of patients reported that they understood the personal implications of the results, whereas 26% did not. The result letter raised questions for 46% of patients and was missing information according to 37%. Questions that were raised remained unanswered for 31% of patients (n=42). Patients wanted to know the exact implications of the results for them, such as the level of dose adjustment (n=12) and what action should be taken (n=12), missed a specific drug being mentioned (n=2), wanted to know more about additional testing and non-genetic factors influencing drug response (n=2), did not understand the appendix meant for HCPs (n=2), wanted to know which HCPs receives the results (n=2), and had questions about the design of and continuation after the study (n=2). Subjects that should be additionally covered in the result letter included the exact implications of the results for the individual patient (i.e. level of dose adjustment) (n=19), explanation of the appendix meant for HCPs (n=9), and what action should be taken (n=5).

After follow-up, 75% of HCPs reported they understood the implications of the results for the care they provide. The result letter raised questions for 35% of HCPs and missed information according to 18% of HCPs, particularly the exact implications of the results for the individual patient such as the level of dose adjustment (n=6) and how the results should be applied (n=4). Questions that were raised during follow-up remained unanswered for 11% of HCPs (n=6), and they wanted to know how to record PGx screening results correctly in their EHRs (n=3) and wanted explanations for the abbreviations (n=1).

These findings indicate that the result letter sent out during this study was not meeting the informational needs of patients and HCPs. Furthermore, HCPs seem to need more practical pointers on how to deal with PGx screening results.
