## Supplementary material for "Practical barriers and facilitators experienced by patients, pharmacists and physicians to the implementation of pharmacogenomic screening in Dutch outpatient hospital care – an explorative pilot study": Table S1

Table S1: Details of custom genotyping panel

| **Gene** | **Reference SNP ID** | **Associated haplotypes** | **Type of variation** |
| --- | --- | --- | --- |
| CYP1A2 | rs2069514 | *1C; *1L | G>A |
|  | rs762551 | *1F; *1L | C>A |
| CYP2B6 | rs3745274 | *6 | G>T |
| CYP2C9 | rs1799853 | *2 | C>T |
|  | rs1057910 | *3 | A>C |
| CYP2C19 | rs12248560 | *4B; *17 | C>T |
|  | rs4986893 | *3 | G>A |
|  | rs4244285 | *2 | G>A |
|  | rs28399504 | *4A; *4B | A>G |
| CYP2D6 | rs1065852 | *4; *10; *36; *69 | G>A |
|  | rs28371725 | *41; *69 | C>T |
|  | rs35742686 | *3 | T>Del |
|  | rs3892097 | *4; *4M | C>T |
|  | rs5030655 | *6 | A>Del |
|  | rs5030656 | *9; *109 | CTT>Del |
|  | rs5030862 | *12 | C>T |
|  | rs5030867 | *7 | T>G |
|  | rs59421388 | *29; *109 | C>T |
|  |  | *5 | Whole gene deletion |
|  |  | *36 | Partial deletion |
| CYP3A4 | rs35599367 | *22 | G>A |
| CYP3A5 | rs776746 | *3 | C>T |
|  | rs10264272 | *6 | C>T |
| Factor V | rs6025 | Leiden mutation | C>T |
| HLA-B | rs2395029 | *5701 | T>G |
| MTHFR | rs1801133 | 677T (high risk allele) | G>A |
| SLCO1B1 | rs4149056 | 521C (high risk allele) | T>C |
| TPMT | rs1800462 | *2 | C>G |
|  | rs1142345 | *3A; *3C | T>C |
|  | rs1800460 | *3A; *3B | C>T |
| UGT1A1 | N/A | *28; *36; *37 | TA-repeat |
| VKORC1 | rs9934438 | *2 | G>A |
