## Supplementary material for "Practical barriers and facilitators experienced by patients, pharmacists and physicians to the implementation of pharmacogenomic screening in Dutch outpatient hospital care – an explorative pilot study": Table S3

Table S3: Demographics of participating patients and healthcare practitioners

| A | Patients (n=165) | Median or n | Range or % |
| --- | --- | --- | --- |
|  | Age (years) | 59 | 19–81 |
|  | Respondents  T1 survey | 138 | 84% |
|  | T2 survey | 122 | 74% |
|  | Gender (male) | 70 | 42% |
|  | T1 respondents  T2 respondents | 59  48 | 43%  39% |
|  | Clinic (internal medicine) | 115 | 70% |
|  | Follow-up |  |  |
|  | Duration (days) | 244 | 117–365 |
|  | Appointments | 3 | 0–162 |
|  | Education level (T1)  Low | 42 | 30% |
|  | Intermediate | 45 | 33% |
|  | High | 51 | 37% |
|  | Drug use management (T1)  Independent | 124 | 90% |
|  | With help | 6 | 4% |
|  | No drug use | 8 | 6% |

| B | Physicians (n=21) | Median or n | Range or % |
| --- | --- | --- | --- |
|  | Age (years) | 49 | 31–76 |
|  | Gender (male) | 10 | 48% |
|  | Clinic (internal medicine) | 14 | 67% |
|  | Working experience (years)* | 6 | 1–46 |
|  | Occupation  Medical specialist | 19 | 90% |
|  | Physician | 2 | 10% |
|  | *in current occupation | | |

| C | Hospital pharmacist (n=13) | Median or n | Range or % |
| --- | --- | --- | --- |
|  | Age (years) | 37 | 25–59 |
|  | Gender (male) | 7 | 54% |
|  | Working experience (years) | 3 | 0–32 |
|  | Occupation  Hospital pharmacist  Hospital pharmacist in training  Pharmacist without specialty | 7  3  3 | 54%  23%  23% |

| D | Community pharmacist (n=48) | Median or n | Range or % |
| --- | --- | --- | --- |
|  | Age (years) | 38 | 25–63 |
|  | Gender (male) | 23 | 48% |
|  | Working experience (years) | 12.5 | 1–37 |
|  | Occupation  Community pharmacist  Community pharmacist in training  Pharmacist without specialty | 44  2  2 | 92%  4%  4% |
