## Supplementary material for "Practical barriers and facilitators experienced by patients, pharmacists and physicians to the implementation of pharmacogenomic screening in Dutch outpatient hospital care – an explorative pilot study": Table S4

**Table S4:** Frequencies of PGx haplotypes and predicted PGx phenotypes in the study population compared to literature

| **Gene**  (number of individuals) ^a^ | **Haplotype / predicted phenotype** | **Frequency (%) in study population** | **Frequency (%) in (Dutch) Caucasian population** | **Reference** |
| --- | --- | --- | --- | --- |
| ***CYP1A2*** | *1A | 29.68 | 24.4 | ^1^ |
| (n = 155) | *1C | 0 | 1 | ^2,3,4^ |
|  | *1F | 69.03 | 67 | ^2,3,4^ |
|  | *1L | 1.29 | 0.8 | ^1^ |
|  | Poor Metaboliser | 0 | - |  |
|  | Intermediate Metaboliser | 0 | - |  |
|  | Normal Metaboliser | 100 | - |  |
| ***CYP2B6*** | *1 | 76.22 | 61.1 | ^5^ |
| (n = 164) | *6 | 23.78 | 3.4 | ^5^ |
|  | Poor Metaboliser | 4.88 | 6–12 | ^6^ |
|  | Intermediate Metaboliser | 37.80 | 15–45 | ^6^ |
|  | Normal Metaboliser | 57.32 | 44–85 | ^6^ |
| ***CYP2C9*** | *1 | 78.05 | 80.01 | ^7^ |
| (n = 164) | *2 | 14.94 | 12.60 | ^7^ |
|  | *3 | 7.01 | 7.08 | ^7^ |
|  | Poor Metaboliser | 3.05 | 4.00 | ^7^ |
|  | Intermediate Metaboliser | 37.80 | 31.99 | ^7^ |
|  | Normal Metaboliser | 59.15 | 64.01 | ^7^ |
| ***CYP2C19*** | *1 | 62.04 | 62.4 | ^8^ |
| (n = 162) | *2 | 17.59 | 14.6 | ^8,9^ |
|  | *3 | 0.31 | 0.02 | ^9^ |
|  | *4A | 0.31 | (*4) 0.28 | ^8^ |
|  | *4B | 0 | (*4) 0.28 | ^8^ |
|  | *17 | 19.75 | 21.3–21.7 | ^8,9^ |
|  | Poor Metaboliser | 4.94 | 2.5 | ^8^ |
|  | Intermediate Metaboliser | 26.54 | 26.8 | ^8^ |
|  | Normal Metaboliser | 64.20 | 66.1 | ^8^ |
|  | Ultra-rapid Metaboliser | 4.32 | 4.6–4.77 | ^8,9^ |
| ***CYP2D6*** | *1 | 48.24 | 41.38 | ^10^ |
| (n = 142) | *1x2 | 0.35 | 0.94 | ^10^ |
|  | *3 | 2.46 | 1.36 | ^10^ |
|  | *4 | 5.28 | 17.93 | ^10^ |
|  | *4x2 | 0.35 | 0.28 | ^10^ |
|  | *4M | 0.70 | - |  |
|  | *5 | 3.52 | 2.65 | ^10^ |
|  | *6 | 0 | 0.92 | ^10^ |
|  | *7 | 0 | 0.07 | ^10^ |
|  | *9 | 1.76 | 2.03 | ^10^ |
|  | *10 ^b^ | 1.06 | 2.32 | ^10^ |
|  | *12 | 0 | 0.01 | ^10^ |
|  | *29 | 0 | 0.16 | ^10^ |
|  | *36 | 0 | 0 | ^10^ |
|  | *41 ^b^ | 0 | 8.88 | ^10^ |
|  | *69 | 0 | - |  |
|  | *109 | 0 | - |  |
|  | Not conclusive (*1/*4 or *4M/*10) ^c^ | 27.46 | - |  |
|  | Not conclusive (*4/*41 or *4M/*69) ^c^ | 2.11 | - |  |
|  | Not conclusive (*1/*10/*36) ^d^ | 0.70 | - |  |
|  | Poor Metaboliser | 7.75 | 5.4–9 | ^11^ |
|  | Intermediate Metaboliser | 40.14 | 10–40 | ^11^ |
|  | Normal Metaboliser | 51.41 | 80 | ^11^ |
|  | Ultra-rapid Metaboliser | 0.70 | 1–2 | ^11^ |
| ***CYP3A4*** | *1 | 92.38 | 91.5 | ^5^ |
| (n = 164) | *22 | 7.62 | 5.0 | ^5^ |
|  | Poor Metaboliser | 0.61 | 0 | ^12^ |
|  | Intermediate Metaboliser | 14.02 | 6.40 | ^12^ |
|  | Normal Metaboliser | 85.37 | 93.60 | ^12^ |
| ***CYP3A5*** | *1 | 8.54 | - |  |
| (n = 164) | *3 | 91.46 | 91.7 | ^13^ |
|  | *6 | 0 | - |  |
|  | Non-expressor | 84.15 | 81.4 | ^13^ |
|  | Heterozygous expressor | 14.63 | 18.4 | ^13^ |
|  | Homozygous expressor | 1.22 | 0.2 | ^13^ |
| ***Factor V*** | Wildtype allele | 94.82 | 95.6 | ^14^ |
| (n = 164) | Leiden mutation | 5.18 | 4.4 | ^14^ |
|  | Homozygous wildtype | 89.63 | 97.05 | ^15^ |
|  | Heterozygous Leiden mutation | 10.37 | 2.9 | ^15^ |
|  | Homozygous Leiden mutation | 0 | 0 | ^15^ |
| ***HLA-B*** | Wildtype allele | 97.53 | - |  |
| (n = 162) | *5701 | 2.47 | 3.4 | ^16^ |
|  | No risk | 95.06 | - |  |
|  | Intermediate | 4.94 | - |  |
|  | High risk | 0 | - |  |
| ***MTHFR*** | 677C (wildtype) | 68.90 | 81.4 | ^17^ |
| (n = 164) | 677T (risk allele) | 31.10 | 18.6 | ^17^ |
|  | No risk | 45.73 | 51.70 | ^18^ |
|  | Intermediate risk | 46.34 | 40.25 | ^18^ |
|  | High risk | 7.93 | 8.05 | ^18^ |
| ***SLCO1B1*** | 521T (wildtype) | 83.94 | 81 | ^19^ |
| (n = 165) | 521C (risk allele) | 16.06 | 19 | ^19^ |
|  | No risk | 70.30 | 68.67 | ^19^ |
|  | Intermediate risk | 27.27 | 24.10 | ^19^ |
|  | High risk | 2.42 | 7.23 | ^19^ |
| ***TPMT*** *^e^* | *1 | 95.12 | - |  |
| (n = 164) | *2 | 0.30 | 0.4 | ^20^ |
|  | *3A | 0 | 3.5 | ^20^ |
|  | *3B | 0 | 0.4 | ^20^ |
|  | *3C | 4.57 | 0.8 | ^20^ |
|  | Poor Metaboliser | 0 | 0.26 | ^20^ |
|  | Intermediate Metaboliser | 9.76 | 10 | ^20^ |
|  | Normal Metaboliser | 90.24 | 90 | ^20^ |
| ***UGT1A1*** | *1 | 67.08 | 61.3 | ^21^ |
| (n = 161) | *28 | 32.30 | 38.7 | ^21^ |
|  | *36 | 0.62 | 0 | ^21^ |
|  | *37 | 0 | 0 | ^21^ |
|  | Poor Metaboliser | 9.94 | 9 | ^22^ |
|  | Intermediate Metaboliser | 44.10 | 54 | ^22^ |
|  | Normal Metaboliser | 45.96 | 37 | ^22^ |
| ***VKORC1*** | *1 (wildtype) | 60.98 | 58 | ^23^ |
| (n = 164) | *2 (risk allele) | 39.02 | 37 | ^23^ |
|  | No risk | 40.24 | 36.3 | ^24^ |
|  | Intermediate risk | 41.46 | 48.7 | ^24^ |
|  | High risk | 18.29 | 15.0 | ^24^ |

^a^ Number of genotyped individuals that passed QC thresholds.

^b^ It was not possible to discriminate between *10, *14, *37, *47, *49, *52, *54, *56B, *57, *65, *72, *87, *94, *95, *100 and *101. The haplotype reported was *10. It was not possible to discriminate between *41 and *91. The haplotype reported was *41.

^c^ It was not possible to discriminate between *1/*4 and *4M/*10, and *4/*41 and *4M/*69. For these diplotypes only, the predicted phenotypes were reported.

^d^ It was not possible to discriminate which star alleles were located in tandem configuration. For this individual only, the predicted phenotype was reported.

^e^ It was not possible to discriminate between *1/*3A and *3B/*3C. The diplotype reported was *1/*3A.

1.	Ghotbi, R. *et al.* Comparisons of CYP1A2 genetic polymorphisms, enzyme activity and the genotype-phenotype relationship in Swedes and Koreans. *Eur. J. Clin. Pharmacol.* **63**, 537–546 (2007).

2.	Zhou, S.-F., Wang, B., Yang, L.-P. & Liu, J.-P. Structure, function, regulation and polymorphism and the clinical significance of human cytochrome P450 1A2. *Drug Metab. Rev.* **42**, 268–354 (2010).

3.	Weide, J. van der, Steijns, L. S. & Weelden, M. J. van The effect of smoking and cytochrome P450 CYP1A2 genetic polymorphism on clozapine clearance and dose requirement. *Pharmacogenetics* **13**, 169–172 (2003).

4.	Söderberg, M. M., Haslemo, T., Molden, E. & Dahl, M.-L. Influence of CYP1A1/CYP1A2 and AHR polymorphisms on systemic olanzapine exposure. *Pharmacogenet. Genomics* **23**, 279–285 (2013).

5.	Zhou, Y., Ingelman-Sundberg, M. & Lauschke, V. Worldwide Distribution of Cytochrome P450 Alleles: A Meta-analysis of Population-scale Sequencing Projects. *Clin. Pharmacol. Ther.* **102**, 688–700 (2017).

6.	Koninklijke Nederlandse Maatschappij ter bevordering der Pharmacie Algemene achtergrondtekst Farmacogenetica - CYP2B6. (2017).

7.	Caudle, K. E. *et al.* Clinical Pharmacogenetics Implementation Consortium Guidelines for CYP2C9 and HLA-B Genotypes and Phenytoin Dosing. *Clin. Pharmacol. Ther.* **96**, 542–548 (2014).

8.	Scott, S. A. *et al.* Clinical Pharmacogenetics Implementation Consortium Guidelines for CYP2C19 Genotype and Clopidogrel Therapy: 2013 Update. *Clin. Pharmacol. Ther.* **94**, 317–323 (2013).

9.	Ionova, Y. *et al.* CYP2C19 Allele Frequencies in Over 2.2 Million Direct‐to‐Consumer Genetics Research Participants and the Potential Implication for Prescriptions in a Large Health System. *Clin. Transl. Sci.* cts.12830 (2020).doi:10.1111/cts.12830

10.	Whirl-Carrillo, M. *et al.* Pharmacogenomics Knowledge for Personalized Medicine. *Clin. Pharmacol. Ther.* **92**, 414–417 (2012).

11.	Koninklijke Nederlandse Maatschappij ter bevordering der Pharmacie Algemene achtergrondtekst Farmacogenetica - CYP2D6. (2020).

12.	Elens, L. *et al.* The new CYP3A4 intron 6 C>T polymorphism (CYP3A4*22) is associated with an increased risk of delayed graft function and worse renal function in cyclosporine-treated kidney transplant patients: *Pharmacogenet. Genomics* 1 (2012).doi:10.1097/FPC.0b013e328351f3c1

13.	Schaik, R. H. N. van, Heiden, I. P. van der, Anker, J. N. van den & Lindemans, J. CYP3A5 variant allele frequencies in Dutch Caucasians. *Clin. Chem.* **48**, 1668–1671 (2002).

14.	Rees, D. C., Cox, M. & Clegg, J. B. World distribution of factor V Leiden. *The Lancet* **346**, 1133–1134 (1995).

15.	Rosendaal, F. R., Koster, T., Vandenbroucke, J. P. & Reitsma, P. H. High risk of thrombosis in patients homozygous for factor V Leiden (activated protein C resistance). *Blood* **85**, 1504–1508 (1995).

16.	Gonzalez-Galarza, F. F. *et al.* Allele frequency net database (AFND) 2020 update: gold-standard data classification, open access genotype data and new query tools. *Nucleic Acids Res.* gkz1029 (2019).doi:10.1093/nar/gkz1029

17.	Schneider, J. A., Rees, D. C., Liu, Y.-T. & Clegg, J. B. Worldwide Distribution of a Common Methylenetetrahydrofolate Reductase Mutation. *Am. J. Hum. Genet.* **62**, 1258–1260 (1998).

18.	Ede, A. E. van *et al.* The C677T mutation in the methylenetetrahydrofolate reductase gene: a genetic risk factor for methotrexate-related elevation of liver enzymes in rheumatoid arthritis patients. *Arthritis Rheum.* **44**, 2525–2530 (2001).

19.	Brunham, L. R. *et al.* Differential effect of the rs4149056 variant in SLCO1B1 on myopathy associated with simvastatin and atorvastatin. *Pharmacogenomics J.* **12**, 233–237 (2012).

20.	Koninklijke Nederlandse Maatschappij ter bevordering der Pharmacie Algemene achtergrondtekst Farmacogenetica - Thiopurine-S-methyltransferase (TPMT). (2019).

21.	Beutler, E., Gelbart, T. & Demina, A. Racial variability in the UDP-glucuronosyltransferase 1 (UGT1A1) promoter: A balanced polymorphism for regulation of bilirubin metabolism? *Proc. Natl. Acad. Sci.* **95**, 8170–8174 (1998).

22.	Koninklijke Nederlandse Maatschappij ter bevordering der Pharmacie Algemene achtergrondtekst Farmacogenetica - UGT1A1. (2014).

23.	Rieder, M. J. *et al.* Effect of *VKORC1* Haplotypes on Transcriptional Regulation and Warfarin Dose. *N. Engl. J. Med.* **352**, 2285–2293 (2005).

24.	Teichert, M. *et al.* Genotypes Associated With Reduced Activity of VKORC1 and CYP2C9 and Their Modification of Acenocoumarol Anticoagulation During the Initial Treatment Period. *Clin. Pharmacol. Ther.* **85**, 379–386 (2009).
